# Effect of rural trauma team development on outcomes of motorcycle related injuries: A protocol for a multi-center cluster randomized controlled clinical trial (The MOTOR trial)

**DOI:** 10.1101/2023.12.07.23299662

**Authors:** Herman Lule, Micheal A. Mugerwa, Robinson SSebuufu, Patrick Kyamanywa, Till Bärnighausen, Jussi P. Posti, Michael Lowery Wilson

## Abstract

**Background:** Injury is a global health concern whose mortality disproportionately impact low-income countries. Compelling evidence from high-income countries show that rural trauma team development courses (RTTDC) increase clinicians’ knowledge. There is a dearth of evidence from controlled clinical trials to demonstrate the effect of RTTDC on process and patient outcomes. We document a protocol for a multi-center cluster randomized controlled clinical trial which aims to examine the impact of RTTDC on process and patient outcomes of motorcycle-related injuries.

**Methods:** This will be a two-armed parallel multiple period cluster randomized controlled clinical trial in Uganda, where rural trauma team development training is not routinely conducted. We shall recruit regional referral hospitals and include road traffic injured patients, interns, medical trainees, and road traffic law enforcement professionals who serve as trauma care frontliners. Three hospitals will be cluster randomized to RTTDC (intervention group) and the remaining three to standard care (control group). The primary outcomes will be prehospital interval from accident scene to arrival at emergency department, and referral-exit interval from the time the referral decision is made to hospital exist in hours as a measure of process improvement. The secondary outcomes will be all cause mortality, and morbidity of neurological, and orthopedic injuries based on the Glasgow outcome scale and trauma outcome measure scores respectively at 90-days post injury. All outcomes will be measured as final values. We shall compare baseline characteristics and outcomes both at individual, and at cluster level as intervention versus control group. We shall use the mixed effects regression models in Stata 15.0 to report any absolute or relative differences along with 95% CIs. We shall perform subgroup analyses to control for confounding due to injury mechanisms and severity. In parallel to the trial, we shall establish a motorcycle trauma outcome registry (MOTOR) in consultation with community traffic police.

**Discussion:** Our results could inform the design, implementation, and scalability of future rural trauma teams and education programs.

**Trial registration:** Retrospectively registered with Pan African Clinical Trial Registry (PACTR202308851460352) on 17 August 2023.

**Ethics and dissemination:** Ethical approval was obtained from Uganda National Council for Science and Technology (Ref: SS 5082) prior recruitment. The findings, anonymized datasets and code for analysis will be published publicly.

**Protocol Version:** V (2023).

**Administrative information:** HL (principal investigator, and corresponding author) was enhanced with a personal research loan from Uganda Medical Association to support this study. JPP is supported by the Academy of Finland (grant no 17379) and the Maire Taponen Foundation. The study sponsors did not have any role in the design, writing or decision to submit the protocol for publication. MAM is the study biostatistician, RS and PK are onsite study overseers whereas TB, JPP and MLW are offsite study supervisors.

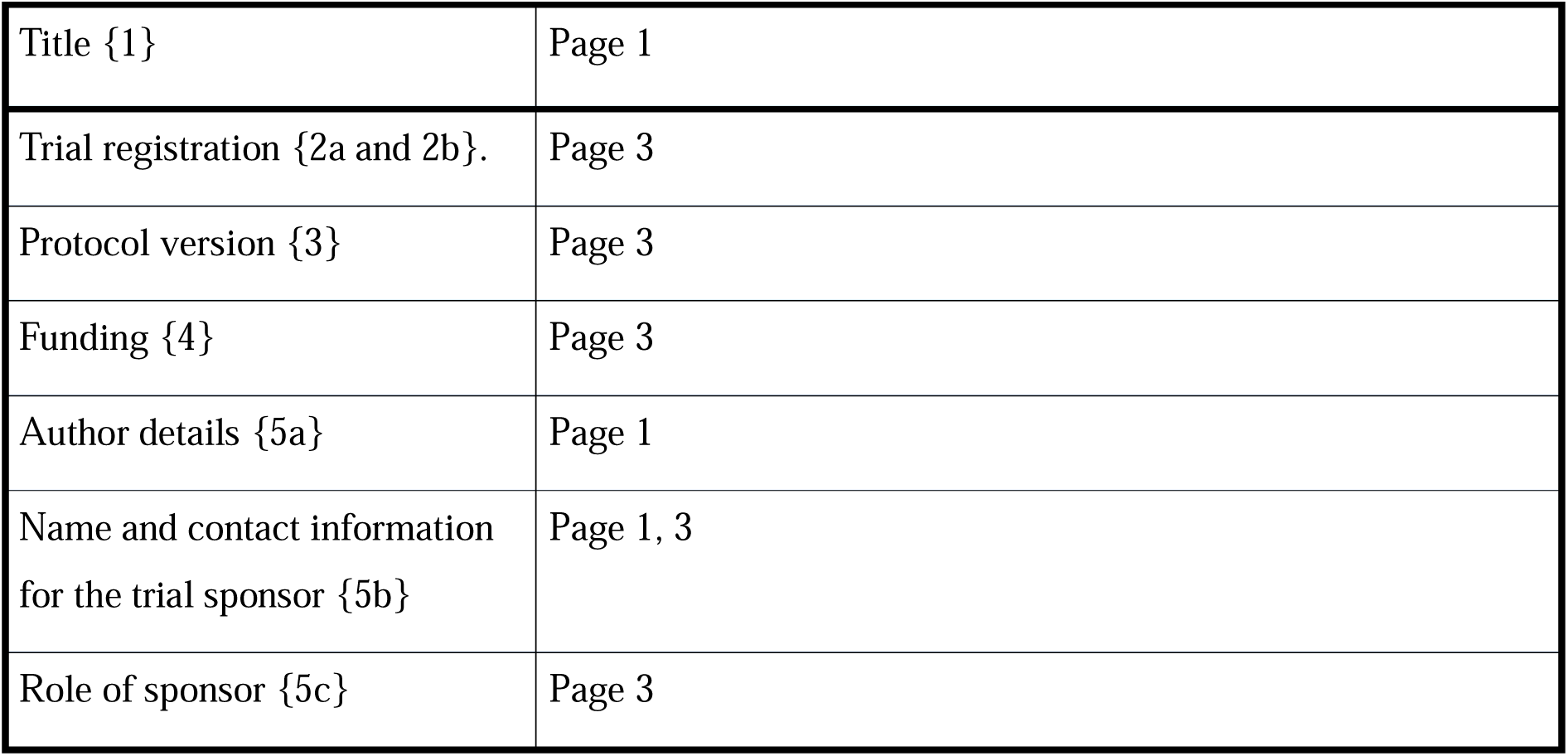

## Introduction

### Background and rationale {6a}

Transport-related injuries are a global public health threat, anticipated to rank fifth in the global burden of disease by 2030 [1]. Evidence suggests that 90% of transport-related crash mortality burden disproportionately occur in low- and middle-income countries (LMICs) such as those in Africa [1], despite concerns of under-reporting [2]. The main cause of injury-related mortality in Africa are road traffic crashes [3] which have serious economic consequences [4]. Uganda is one of the LMICs in Africa with a high road traffic injury burden [5]. Our recent studies show that most of the fatal injuries in Uganda result from motorized two wheel-car collisions, contributing to (52.6%-64.7%) of orthopedic and traumatic brain injuries [6], [7], [8]. Further, the majority (74%) of Uganda’s population live in rural areas [9], which could partly explain its high injury-related mortality rates. Research show that the risk of trauma-related deaths in rural areas increases with remoteness due to health care disparities such as: large commute distances, scarce resources, delays in referrals, lack of skilled trauma care providers as well as population specific and contextual challenges such as poverty and impassable road networks [10].

Uganda lacks adequate universal access to trauma care and injury surveillance information systems due to limited human, infrastructure, and social security insurance resources for healthcare funding; thus its sluggish progress towards attaining sustainable development goal (SDG) III [11]. As such, in terms of level of specialty care, the ratio of surgeon: patient is 0.7: 25,000, notwithstanding the fact that interns and medical trainees contribute to more than 75% of the Uganda’s health workforce [12], [13]. Research show than more than half of the orthopedic and neuro trauma patients in Uganda do not receive first aid prior hospital arrival [6], [7]. Moreover, most of such patients arrive to their definitive trauma centers far beyond the recommended golden hour, either by public means such as motorcycles [8] or by police vans, without addressing the key determinants of trauma-related mortality such as blood pressure and oxygen circulation in transit [7], [14]. These factors have threefold implications. First, the need to strengthen capacity amongst lay responders. Secondly, the need to stabilize patients at rural regional trauma centers prior transfer, and lastly ensuring closed loop communication between lower (level 3) and higher (level 1) trauma centers. The key challenge to address in Ugandan rural settings is the lack of a formal pre-hospital care system as evidenced by emergency evacuation and transfer of trauma patients by untrained police [14]. Further, there is immense need to redress the weak immediate post-crash care response, it being that medical trainees are the first point of post-crash contact as opposed to specialists [12]. We visualize fixing these challenges at community level, and at level 3 rural trauma centers by creating and providing training capacity for rural trauma networks between traffic police and medical interns/trainees at regional referral hospitals.

### Rationale

Evidence from two systematic reviews show that trauma education programs have potential for skills acquisition and knowledge retention amongst health professionals, however for the case of LMICs, most of such programs have not been locally contextualized, and have been largely assessed based on their theoretical merit rather than their clinical impact on patients [15], [16]. A recent observational study in Portugal showed that the European trauma course (ETC) improved self-efficacy and organization skills in individual routine practice but demanded that future investigations should be conducted to examine the effect of the training on trauma outcomes [17]. This European study together with systematic reviews in LMICs have recommended high-caliber epidemiological studies to evaluate the effect of such educational programs on process outcomes such as organizational efficiency, and on patient outcomes such as morbidity and mortality [15], [16], [18]. Except for the current ongoing trial comparing primary trauma course (PTC) to advanced life support (ATLS) and routine care in India [19], trauma education programs in LMICs have mostly been evaluated based on non-randomized studies which has limited their clinical uptake [15]. Moreover most of the programs limit the course participants to qualified hospital-based medical providers [18], while interns, medical and allied health trainees, and road traffic law enforcement professionals are the most readily available frontliners for injured patients in LMICs [14]. Inclusive planning while leveraging on services that have no trauma designation such as police is critical for expanding and strengthening complex rural trauma systems to improve injury reporting [2]. Locally contextualized trauma training has been identified as one of the most crucial steps to operationalizing nonfunctional rural trauma networks [10]. However in a scoping review by Brown et al [18], only twelve out of thirty four trauma courses in LMICs had been contextualized to suite low-income settings. Moreover, a cautiously executed integrative literature review on challenges faced by trauma systems recommend that the next step for future research should be an examination of how trauma training impacts outcomes of patients in remote environment [10]. This multi-center cluster randomized controlled trial will address this gap by examining the effect of a locally contextualized rural trauma team development course (RTTDC) of the American College of Surgeons [20] on process and patient outcome measures in Ugandan trauma centers. The trial will focus on musculoskeletal and neurological injuries which have been recently identified as the most important targets for strengthening health systems in rural Uganda [8], and is developed in accordance with the SPIRIT guidelines for reporting outcomes in trial protocols and the numbers in parentheses {} represent individual items in the SPIRIT checklist [21].

### Objectives {7}

#### Broad objective

The main objective of this trial is to determine the effect rural trauma team development course training on the clinical-process time efficiency and patient centered outcomes of motorcycle related injuries.

#### Specific objectives

1. To determine the effect of rural trauma team development course training on crash scene-admission interval and outcome of musculoskeletal injuries.
2. To determine the effect of rural trauma team development course training on the referral decision to hospital exit interval and outcome of neurological injuries.

#### Null hypothesis

The null hypothesis of this trial is that rural trauma team development course training has no effect on the clinical process time efficiency and outcomes of orthopedic and neurological injuries.

### Trial design {8}

This will be a pragmatic two arm parallel cluster randomized, multi-period controlled clinical trial with one intervention (RTTDC) and one control arm (standard care). The trial had been set to start in 2019 but was disrupted due to Covid-19, and the subsequent prioritization of Covid-19 related studies by trial registers and targeted journals led to the delayed publication of this protocol. This trial will embrace simultaneous cluster randomization which is an ideal method for evaluating community level interventions to overcome ethical constraints that make randomization at individual level impractical, while avoiding probable contamination between control and treatment groups [22]. Moreover simultaneous randomization will enable timely operationalization and concurrent participant enrollment since all the outcomes will be measured as final values and there are no foreseen plans for study modifications in accordance with Esserman et al [23]. This arrangement is based on the fact that our previous studies already provided an insight on the likely loss to follow-up rates, baseline mortality rates amongst our targeted trauma population, and informed the feasibility and relevance of collecting the intended primary and secondary outcomes [6], [7].

## Methods: Participants, interventions, and outcomes

### Study setting {9}

This study will be conducted in six specialized teaching and regional referral hospitals in Uganda where RTTDC is not taught, including: Kiryandongo, Jinja, Hoima, Fort portal, Mubende and Kampala International University Hospital. These facilities have similar characteristics by serving as teaching, residency and internship sites for undergraduate, graduate medical doctors and nurses. The hospitals offer 24/7 emergency surgical services for trauma patients through multidisciplinary teams of orthopedic, general, and visiting neurosurgeons as well as imaging, rehabilitation, and physiotherapy. Each surgical department in these facilities is typically composed of about 1-4 faculty, 2-4 specialty residents often referred to as senior house officers (SHOs), 4-6 interns and 10-30 undergraduate medical trainees. These facilities are suited for this study since they serve both rural population and newly created populous cities that are still struggling with urban planning. Uganda is the 8^th^ and 30^th^ most populous country in Africa and worldwide respectively [9], and its trauma surveillance systems are in infancy stage with weak pre- and pos-crash response [6], [12]. Despite efforts made in the past decade by Ugandan government to address the unmet need for trauma care through infrastructure development, such as equipping operating theaters and intensive care units in public hospitals; limited standardized pre- and in-service trauma training, pre-hospital delays and human resource constraints still contribute to preventable trauma mortalities [14]. To guarantee the quality of data for this project, a prospective motorcycle trauma outcome registry (MOTOR) will be piloted at the participating six regional referral hospitals in parallel to this trial.

### Eligibility criteria {10}

Inclusion criteria

a. Study sites: Level 3 trauma centers which are teaching hospitals, staffed with medical trainees, interns, surgery residents and consultants; offering 24/7 emergency surgical care with access to blood banks, ultrasound, X-ray, and CT scan as locally available or as outsourced services.
b. Trainee participants: Third year or fifth year medical or allied health students, interns or specialty residents in surgery and traumatology clinical rotations who will stay at the hospital of attachment for at least two-three months. In addition, we shall include road traffic law enforcement professionals concerned with evacuation of trauma patients from accident scenes.
c. Patient participants: Patients who sustain motorcycle-related injuries and present to study sites within 24 hours following crash as defined in previous studies [6]. These will include passengers on motorcycles, motorcycle riders, pedestrians hit by motorcycles, patient suffering from of motorcycle-motorcycle collisions or motorcycle-static object collisions, passengers on a motorcycle or cyclists suffering from motorcycle-car collisions.

#### Exclusion criteria

a. Trauma centers which do not offer placements and teaching facilities for students, interns, and residents.
b. Trainee participants: Medical students who have not commenced their surgery clinical rotation and those not directly involved with care of trauma patients at the time of the training since the trainee medical participants are expected to have already been introduced to surgery, emergency trauma resuscitation concepts and trauma clinical scenarios through clerkships onto which we shall build the rural trauma team concept.
c. Patient participants: Pregnant women, neonates, and infants 0-23 months will be excluded due to the known teratogenic effects of radiations to this population since trauma evaluation in this study will involve obtaining radiographs for orthopedic injuries and Computerized Tomographic (CT) scans for neurological injuries. Patients with documented stroke will also be excluded since the study outcomes involve assessment for functional, physical, and neurological disability directly attributable to trauma. Mentally incapacitated patients who have no legally authorized representatives to sign an informed consent will be excluded as well as patients who die before hospital arrival or before imaging results are obtained. Patients who are passengers in a car or are drivers in a car at the time of crash will be excluded given the “protective casing of the car body” which would make these patients less vulnerable to direct impacts compared to pedestrians, cyclists, or passengers on motorcycles. In addition, elderly over 80 years will be excluded due to their increased risk of fragility fractures that could misrepresent the severity of trauma.

### Who will take informed consent? {26a}

Written informed consent will be obtained for both trainee and patient participants prior to participation. The participants or their legally authorized representatives (for minors and unconscious) will endorse a predesignated consent form document with their signatures in the presence of principal investigator or research assistants (surgery specialty residents). The official informed consent form documents for Mbarara University of Science and Technology (MUST) will be adopted for this purpose. Both trainee and patient participant informed consent documents are available as online supplementary materials 1 and 2 respectively. The ethical committees ruled that trainees involved in data collection process at control centers do not need to provide any consent since this study will neither directly affect their practices nor collect their personal data.

### Additional consent provisions for collection and use of participant data and biological specimens {26b}

There will be no biological specimens retained for this study. The informed consent form documents will have provisions for consenting to follow-up, use of data for approved ancillary studies and for permission to archive the anonymized data on a public data repository.

## Interventions

### Explanation for the choice of comparators {6b}

The **intervention** for this trial is the fourth edition of rural trauma team development course (RTTDC) of the American College of Surgeons [20], which will be delivered to medical trainees, interns, and traffic law enforcement professionals at the three intervention study centers (intervention group). Specialty surgical residents will be trained as faculty who later will serve as trainers since they directly supervise interns and undergraduate medical students in Ugandan settings. The research and ethics committees approved training residents as opposed to specialists to avoid constraining the scarce specialized human resource since this is a trainee-capacity building trauma education program. Moreover, consultants rarely attend to trauma patients for immediate resuscitation as the first point of contact in Ugandan settings. The patient participants at intervention sites will be those with motorcycle-related injuries at any of the study sites. The comparators in this trial are the control group of hospitals which will offer standard care to eligible patients, without their care providers receiving RTTDC training intervention.

### Intervention description {11a}

The 2 day RTTDC interventional training will be conducted in designated spacious multimedia surgical simulation conference rooms at the study sites randomized to intervention arm, and will be delivered in its described standard format [20]. This training model was the most appropriate for Uganda, with its health worker: patient ratio of 1:25,000 serving its 48 million population [12]. The model dwells on team concept to improve coordination and efficiency of the existing “skeletal” health structure in rural environment. We hope to train a total of 66 road traffic police officers, 12 specialty residents, 30 intern doctors, 140 fifth year and 264 third year medical students. These figures were determined based on the average annual number of traffic law enforcement and trainees received at the respective regional police headquarters and hospitals respectively as detailed here [24]. Our target is to train at least 80% of each cohort of eligible trainees received every after 1-5 months within the surgery department during the four year study period until the required trainee and patient participant sample sizes are attained. If trainees drop out prior to completion of data collection, compensations will be made to maintain this criterion during the next cohort of surgical rotation trainee intake. The specifics of how the training will be conducted is detailed elsewhere [24]. The research core team agreed that trainee participants who will score more than 60% in post-training trauma based MCQs at intervention sites will be retained to form a rural trauma team network which will constitute multiple local “ six-team member trauma committees of first responders” with defined roles i.e., each team of five medical trainees having one contact road traffic officer.

To enable an active “alarm” activation criterion for the trauma committees, closed loop communication and smooth “handover” processes from police to medical trainees prior patient transfer, the trained traffic officers on highways will serve as focal contact persons for the rural trauma teams, beginning at parish (sub-county) level. Teammates will freely contact each other in the event of receiving a case of road traffic crash, both within and across teams to coordinate evacuation, consultations, referrals, and arrivals at recipient centers, with support of trained interns and surgery residents as team leaders. Weekly “audit meetings” will be conducted remotely via zoom to discuss team challenges, otherwise a trained surgery resident will be available 24/7 to advise on referrals, transfers, and treatments in liaison with consultants who may be physically or remotely available. The control hospitals in this trial will be left to provide their routine standard care without RTTDC intervention as summarized in Fig. 1.

**Fig. 1:**
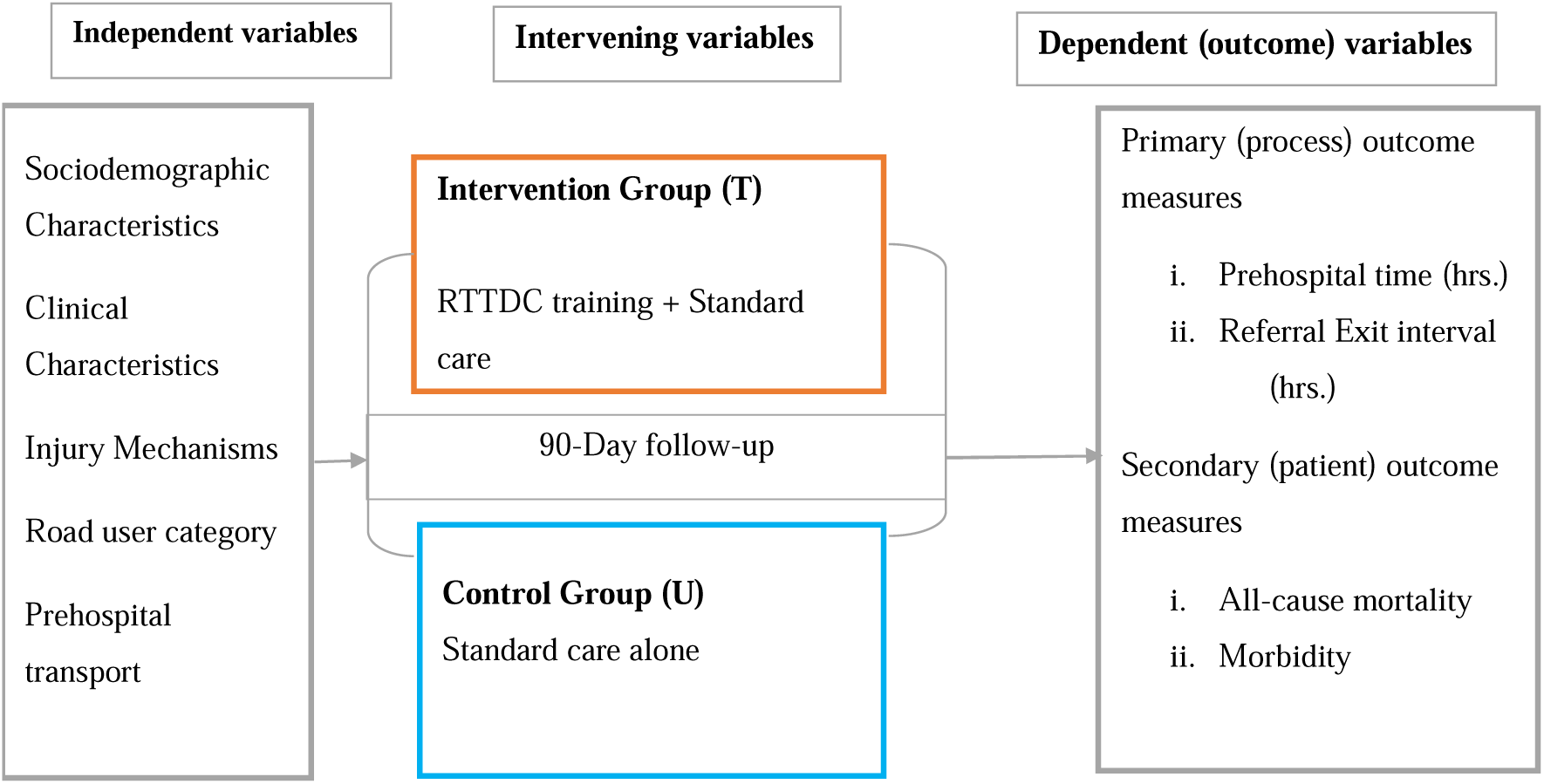
Conceptual framework summarizing interventions and interaction of variables.

### Criteria for discontinuing or modifying allocated interventions {11b}

The 4^th^ edition of RTTDC will be delivered in its standardized form [20] without any modifications.

### Strategies to improve adherence to interventions {11c}

RTTDC is the intervention thus participants who will not be able to attend all the modules and complete the post training MCQs will be excluded. Adherence to RTTDC teachings amongst care providers during their clinical practice such as time taken from referral decision to execution and improper closed loop communication leading to loss to follow-up will be assessed as process measure outcomes.

### Relevant concomitant care permitted or prohibited during the trial {11d}

The baseline quality of care given to trauma patients at each of the participating six hospitals is based on the regulations of the Uganda Medical and Dental Practitioners Council (UMDPC) which regulates medical practice and the National Council for Higher Education (NCHE) which regulates both undergraduate and graduate surgical curricula [25]. The councils require that graduating doctors undergo clinical rotations in surgery and traumatology, clerk, present and document trauma scenarios in patients’ case files. In addition, medical graduates should demonstrate competency in initiating emergency resuscitation in accordance with the advanced trauma life support protocols [26], such as execution of primary and secondary surveys, carefully discern, request and comprehend the necessary imaging and laboratory investigations in trauma, acknowledges self-knowledge and resource limitations and consults or refers patients whose surgical care demands exceed own skills or local trauma capacity. Further, upon graduation, Ugandan doctors would have to obtain a minimum of 48 hours of continuous professional development (CPD) points annually to maintain their licensure.

In terms of standards of care, both intervention and control facilities have medical trainees, interns, specialty residents and specialists who provide care for injured patients. The care typically involves receiving trauma patients who may be brought by staffed ambulances, police or public means to the accident and emergency units, and are received by trainees, interns, or surgical residents. These cadres open case files, initiate immediate care through horizontal consultations, request X-rays, CT scans and laboratory workup; plan definitive care or referrals in liaison with a senior surgical resident, and in consultation with a multidisciplinary team of specialists on call. As such, procedures such as surgical suturing and toileting, limb fracture casting and immobilization, chest tube insertions are often a business of interns. The more demanding splenectomies, external fixation of fractures and burr holes are a matter of specialty residents after notification of a consultant on duty whereas a major case of poly trauma would warranty the decision of a consultant specialist on call before the patient is taken to the operating room. The imaging and blood transfusion services are often freely available at the emergence units as government services but may be outsourced from private hospitals at a cost due to long waiting cues or when service maintenance such as for CT scanners and X-ray machines is underway. Finally, in terms of patient disposition, minor injuries that require surgery under local anesthesia are discharged on the same day, major trauma cases that require conservative treatment such as closed fractures are treated on surgical wards whereas those deeply unconscious or require critical care after major operations are admitted to the intensive care units (ICU) until they are fit for the care on general surgical wards prior discharge. Alternatively due to overcrowding, often with floor cases; patients with major trauma may be referred out and such common reasons for referral include further neurosurgical evaluation with MRI or by a neurosurgeon, need for multiple specialty care and rehabilitation, or for ICU admission elsewhere when the ICU beds are locally limited.

### Provisions for post-trial care {30}

Any participants requiring further care beyond the 90-day follow-up will be linked to their attending clinicians during an additional three-months period after completion of the trial. There will be no compensation other than transport refunds for participants whose visits will be solely for the purposes of study follow-up outside routine clinical visits since this study was considered minimal risk by the approving research ethics committees. The principal investigator will refund the imaging costs for request approved by attending surgeons and radiologists in the unprecedented event of the services being unavailable free of charge.

### Outcomes {12}

This trial will examine both process measures and patient outcomes. The **primary (process) outcomes** of this trial are to compare in hours (i) the prehospital time interval from accident scene to hospital admission, and (ii) the time interval from referral decision to hospital exit amongst patients with motorcycle-related injuries presenting at the intervention versus control study centers.

The **secondary (patient) outcomes** of this trial are to compare: (i) all-cause mortality at 90 days from time of injury, and (ii) morbidity of motorcycle-related orthopedic and neurological injuries presenting at intervention versus control study centers. The morbidity of orthopedic injuries will be measured based on trauma expectation factor score (TEFS) at admission and trauma outcome measure score (TOMS) at 90 days as reported to outcome assessors [27], whereas for neurological injuries, Glasgow Coma Scale (GCS) at admission and Glasgow Outcome Scale (GOS) at 90 days will be used as documented by the attending clinicians [28].

The **tertiary outcomes** will be the effect of the training on providers knowledge based on pre-and post-training trauma related MCQ scores and the barriers to injury care faced by providers during execution of definitive treatment and these will be determined in two ancillary studies that were approved as part of this trial whose protocols are detailed elsewhere [24]. A summary of all outcomes and their case definitions, specific measurement variables, analysis metrics, methods of aggregation and time points is available as online supplementary material 3.

These patient outcomes were selected for our trauma population based on their validity and relevance in previous studies [7], [29]. Further, level III evidence suggest that prolonged prehospital times may be associated with increased mortality of trauma patients [30] but how this relates to time-sensitive orthopedic and neurological injuries in LMICs which lack formal coordinated prehospital systems is mysterious [31]. Thus the process outcome measure variables were selected within the context of evidence-based practice which recommends the golden hour rule that warranty a major trauma patient seeking definitive care to be in the right place within 60 minutes following injury, otherwise mortality or morbidity may be eminent [32]. These variables align well with the proposed outcome data collection to guide resource allocation amongst vulnerable populations that seek emergency services to improve global surgery framework for national surgical, obstetric and anesthesia care provider plans [33]. In addition, the variables expand on outcomes for trauma informed interventions [34]. All outcomes will be measured as final values but will be discussed in relation to the baseline established in our prior feasibility studies [7], [29]. The comparisons in outcomes shall be made at both individual and cluster levels, using the mixed effects regression models.

### Participant timeline {13}

First, cluster randomization of the trauma centers will be performed three weeks prior commencement of the training to determine which centers will receive the training. Potential participants for the training will be screened for eligibility by hospital and study administrators two weeks prior the training, and eligible trainee participants will be consented and assigned to “rural trauma teams” of six members on the first day of the training, during which they will complete a pre-training questionnaire on trauma based multiple choice questions (MCQs). Subsequently trainee participants will be followed up at 90 days (three months) to complete post-training MCQs. The first eligible patient participants at both control and intervention sites will be enrolled concurrently through a motorcycle trauma outcome registry (MOTOR) that will run parallel to the trial. Informed consent, baseline sociodemographic and clinical characteristics, and pre-hospital intervals will be obtained at admission as well as Glasgow Coma Score (GCS). The trauma expectation factor score (TESF) will be obtained during the first week of admission or prior discharge on assumption that patients would have received their definitive surgical intervention or referred for further care during this period. Further, the Glasgow outcome scale (GOS), all-cause mortality and trauma outcome measure score (TOMS) will be obtained at 90-day follow-up. The GCS assess the level of traumatic brain injury (TBI) in the acute phase whereas the GOS determines neurological outcome within the context of level of independence at a later phase of TBI [28]. On the other hand, TOMS assess patient reported trauma outcomes with reference to their expectations (TEFS) at the time trauma intervention in relation to the level of pain, physical function, disability, injury treatment satisfaction and overall satisfaction [27], [29]. Throughout the study period, any individual patient barriers encountered during the pathway to execution of definitive injury care will be documented as summarized in timeline Fig. 2.

**Fig 2.**
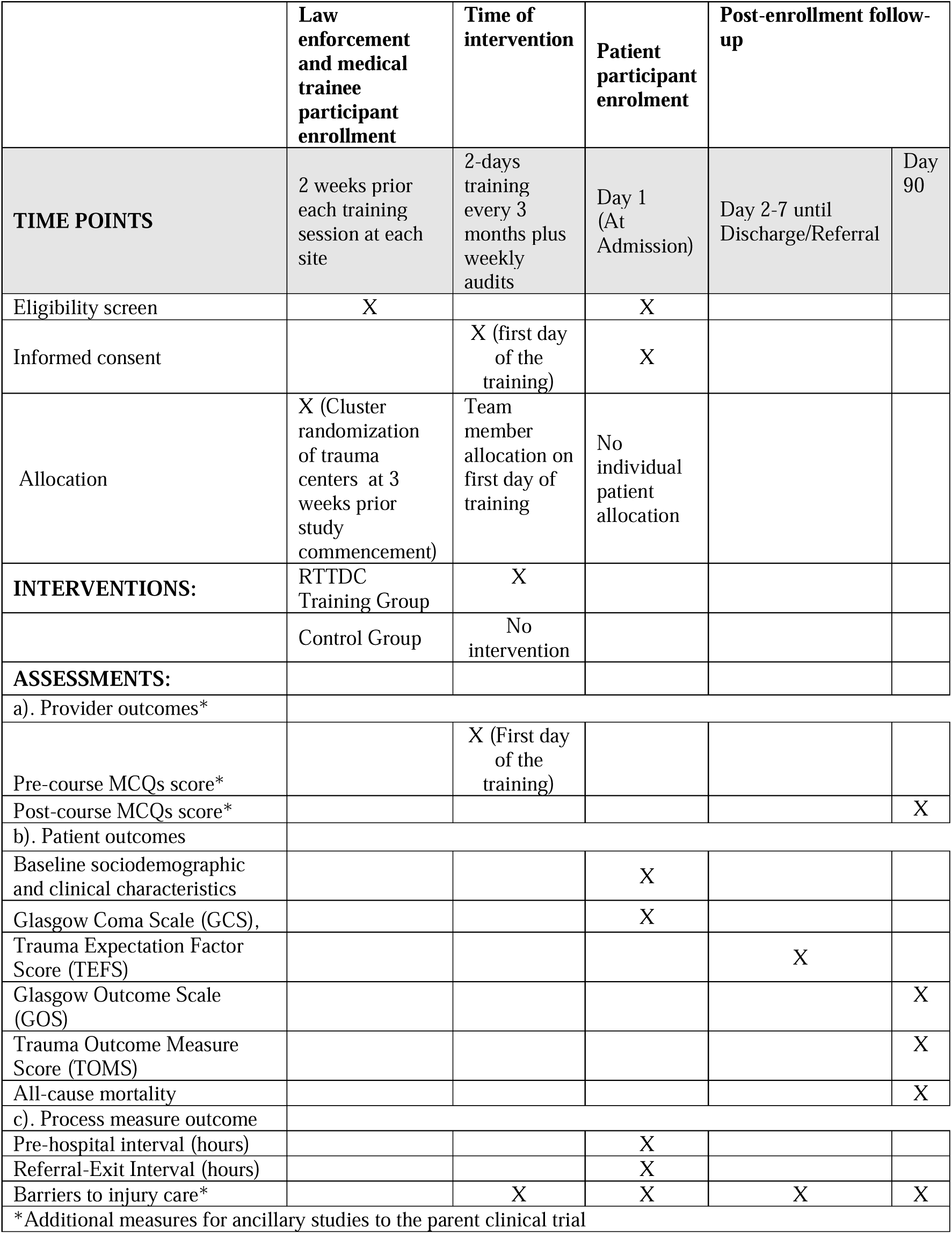
SPIRIT figure showing timelines.

### Sample size {14}

To estimate the sample size, we used the open source R shiny application for cluster randomized controlled trials with parallel design and discrete time decay correlation structures for multiple periods [35] available at https://clusterrcts.shinyapps.io/rshinyapp/. Further, we assumed t-distribution due to plans for small sample corrections at the analysis stage in accordance with Rutterford et al [36]. The study was approved for four years but the core research team foresaw it feasible to actively collect data for a period of 3 years putting into consideration university semester breaks, public holidays, and unexpected events. The three years (36 months) would yield 12 study periods given the planned training of cohorts of medical students on three-monthly basis in accordance with the average duration of internship deployment by the Ugandan ministry of health (1-5 months) and the average duration of surgery clinical rotation for university medical students (2-3 months).

Fixing the study power at 80% for parallel cluster randomized trials with different cross-sections and a discrete time decay period of every three months for a total of 12 periods, and assuming correlations to decay with each period, we explored the minimum number of patient participants (cluster size) required per period using R shiny application [35]. Assuming significance level of 0.05, within period intra-class correlation (ICC) of 0.02 (lower extreme 0.01; upper extreme 0.05), cluster autocorrelation coefficient (CAC) (the ratio of between period ICC to within-period ICC) of 0.8, allowing for varying cluster sizes with a coefficient of variation of 0.5; and based on a mean difference of 1.02 hours in prehospital transfer time and pooled standard deviation of 1.64 hours as continuous outcomes reported in a previous USA based observational study [37]; the upper ICC and base CAC plateaued between 5-10 participants (Fig. 3A). Taking the maximum of 10 participants per cluster period, 80% power could be attained (Fig. 3B); and could be met with a total of three clusters per arm (Fig 3C). This meant a total of 6 study centers (clusters) drawn from a pool of 17 which reasonably represents 35.3% of Uganda’s regional referral hospitals. The sample required per arm is a result of 10 participants per cluster period multiplied by 12 cluster periods multiplied by 3 clusters, yielding 360 participants. The total sample required for two arms would be 720. However, we inflated the sample and variance to cater for the clustering design effect, on assumption that this increases statistical power in accordance with Hemming et al [35]; thus derived sample size (s)= n (1+ (m-1) ρ; where s= required sample size, m =cluster size per period i.e., 10 and p=within period intra-class correlation (0.02), yielding 850 participants.

**Fig. 3A:**
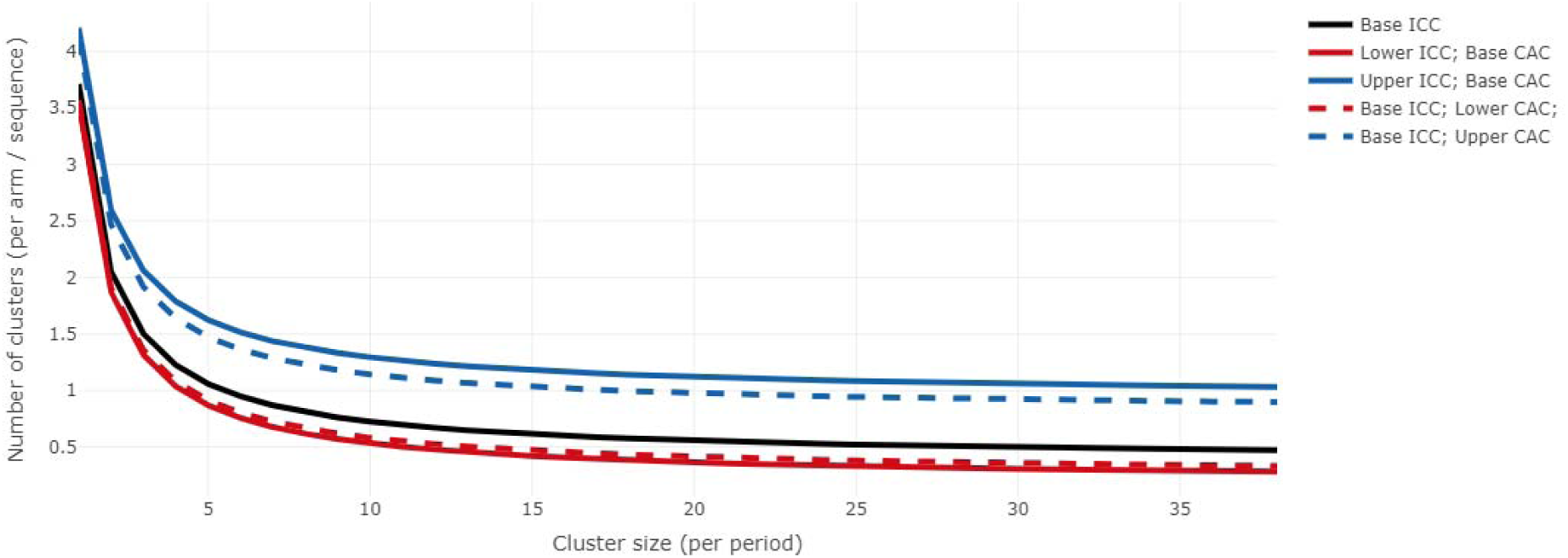
Reduction in number of clusters required as cluster-period size increases at fixed power of 0.8.

**Fig. 3B:**
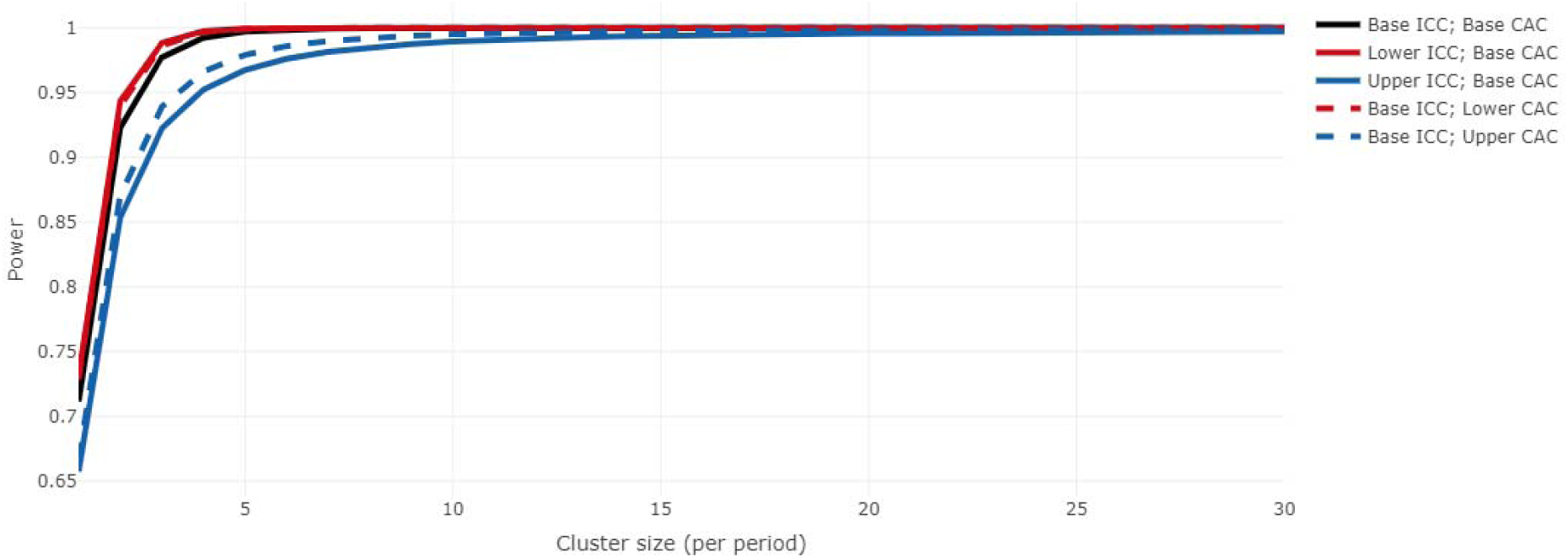
Showing cluster-period size of 10 meeting the 0.8 power requirement for 12 periods.

**Fig. 3C:**
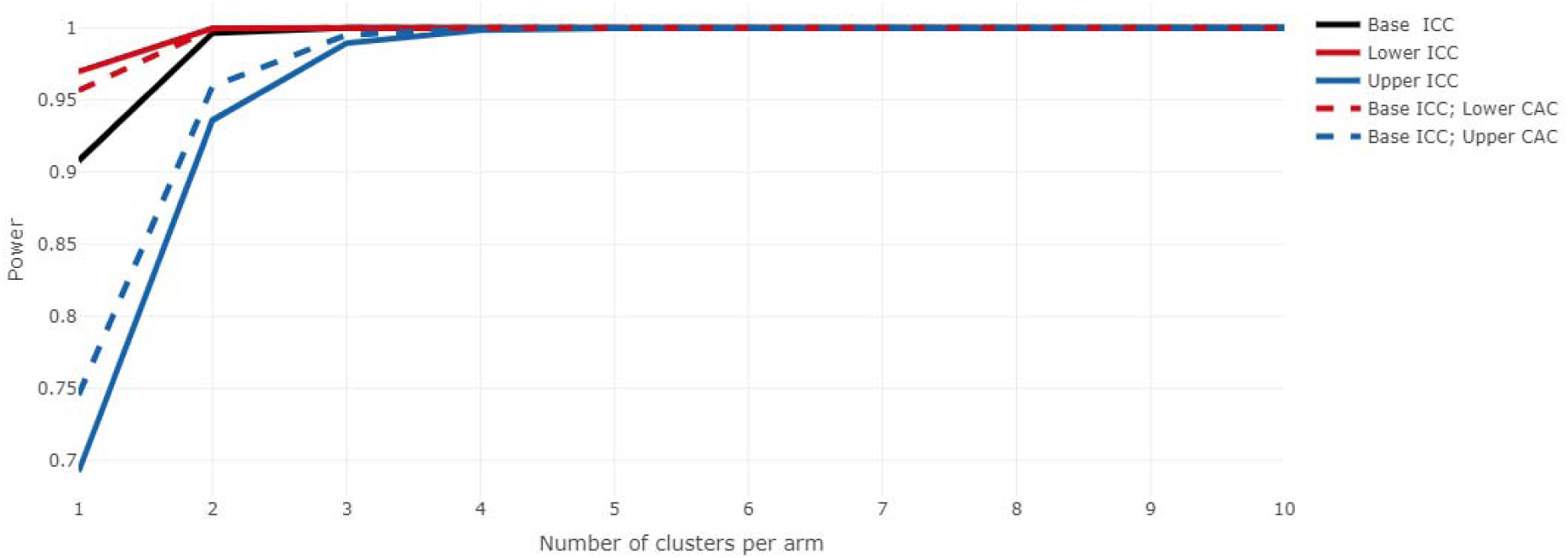
Showing 3 clusters per arm meeting minimum required power of 0.8 for 12 period.

We shall add loss to follow-up rate of 18%, assuming the worst scenario based on our previous dropout rate of 9% in one of the feasibility studies [7]. Therefore, the final sample is approximately 1003 participants. Assuming an equal allocation ratio of 1:1 means 502 per arm and about 168 individuals per cluster. This sample is deemed feasible due to the high trauma burden in Uganda. According to a study at one of the Ugandan rural regional referral hospitals, a total of 900 motorcycle related injuries were recorded during a three year period of which 30.9% (278) and 15.8% (142) sustained musculoskeletal and neurological injuries respectively [8]. We shall not compute sample sizes for each individual secondary outcomes since previous studies did not report any difference in mortality [38].

### Recruitment {15}

The training will be advertised by hospital administrators, class representatives and regional traffic law enforcement leaders and course participants will be recruited by the principal investigator and the respective hospital administrators. The patient participants will be recruited by dedicated project officers at the accident and emergency departments.

## Assignment of interventions: allocation

### Sequence generation {16a}

We shall perform cluster randomization for a permuted block size of six hospitals (clusters) using an open source simulation software [39] available at (https://www.sealedenvelope.com/simple-randomiser/v1/lists), with seed numbers that will be kept confidential to the offsite study administrator. A list length of six random codes will be generated to assign hospitals as intervention or control groups. Since we plan to perform analyses at both individual level and treatment arm levels, we shall not add any stratification factors in the simulation model.

### Concealment mechanism {16b}

The allocation sequence and assignment codes will be generated by and kept secret by an offsite study administrator. Both outcome assessors and patient participants will be blinded by the treatment allocation.

### Implementation {16c}

Trainee participants will be enrolled by the respective university hospital administrators whereas patient participants will be enrolled by project medical officers at the respective study sites who will serve as outcome assessors.

## Assignment of interventions: Blinding

### Who will be blinded {17a}

The trial patient participants and outcome assessors will be blinded using blocked allocation sequence codes.

### Procedure for unblinding if needed {17b}

Unblinding will only be permissible in the likely adverse event to the study participants that can be directly attributable to the study intervention. Such events will be discussed with the attending physicians. Otherwise, the allocation codes will only be revealed to the biostatistician at the time of interim analysis when half of the sample size is recruited. At that point unblinding the biostatistician is considered more beneficial to inform termination or continuation of the trial which outweighs the perceived fear of unknown bias [40].

## Data collection and management

### Plans for assessment and collection of outcomes {18a}

Data collection will start simultaneously at all the study sites. Medical officers (blinded outcome assessors) with a minimum qualification of Bachelor of Medicine and Bachelor of Surgery (MBChB), and at least two years of clinical experience will prospectively collect data through clinical observations, interviews, home visits, and extraction from hospital and police records. All outcome assessors must be willing to undertake a one day budgeted training in the use of data collection tools for this trial and WHO ICD-10 online coding module. The data will be collected daily since medical trainees and interns attend clinical rotations every day. Upon obtaining informed consent, the venue for data collection including follow-up information will be at the accident and emergence departments during admission, surgical wards, out patent clinics, home visits and through phone calls to patients, care takers or recipient hospital physicians in case of referrals.

The study variables will include sociodemographic and clinical such as sex, age, injury mechanisms including nature of collision and road user category, time of injury, prehospital transit time, prehospital care, mode of arrival, vital signs, nature of physical injuries, radiological findings, and injury severity based on GCS and Kampala Trauma Score (KTS) [41]. In addition, data on nature of treatment given, need for referral, referral-exit intervals, and outcomes including TOM, GOS, mortality and time from injury to death will be obtained. The data collection tools used in this trial such as GCS, GOS, TEFS, TOMS, KTS have demonstrated excellent criterion internal validity, consistence and reproducibility in previous studies [41], [27], [28], [42]. Further, the tools are recognized for their higher inter rater reliability, sensitivity to change, and ability to be used as continuous or categorical variables in validation studies [41], [27], [28], [42]. The baseline TEFS, GCS, KTS will be added as covariates whose interaction and confounding will be assessed and controlled since inclusion of baseline values of outcomes as covariates is arguably one of the strongest factors to reduce intraclass correlation coefficient (ICC) estimates [43]. The difference in distribution of baseline characteristics will be compared using means (SD), medians (IQR) and ranges for quantitative data otherwise frequencies and percentages will be used for categorical variables. All data collection forms for this trial and detailed definitions of assessment tools are available in the protocol as supplementary material 4 and 5.

### Plans to promote participant retention and complete follow-up {18b}

Coffee breaks will be facilitated during training to optimize participation. Phone and email contacts will be obtained from all medical trainees and law enforcement professionals. In addition, two phone contacts will be obtained for each patient participant at the time of enrollment at accident and emergency departments i.e., for the patient and their next of kin or legally authorized representative who will in turn be availed with the phone contact of the study nurse coordinators for purposes of follow-up. Automated reminders will be sent to data collection assistants through mobile phones and research electronic data capture (REDCap) platform. Where applicable, home visits will be conducted for participants who are unable to turn up for appointments because of their disability. Trasport refund will be reimbursed for traffic law enforcement professionals and patient participants turning up for appointments outside of the routine hospital visits. Participants will be considered as lost to follow-up if they are untraceable by phone, clinic appointment or home visit. Data on baseline sociodemographic and clinical characteristics of those lost to follow-up or discontinued because of breach of intervention protocols will be retained for comparison between groups but that for withdrawals from consent will be deleted.

## Patient and Public Involvement

One day consultative meeting was conducted at each study site with chief residents, heads of interns, patient care givers, student representatives, and regional traffic police commanders in the month which preceded the study commencement to discuss their perceived barriers to injury care to uncover themes for potential sources of delays ranging from crash scene discovery, evacuation, and prehospital transportation to emergency care which shaped the final data collection tools. During this engagement, trainee and care giver representatives gave an insight on the feasibility of the outcome assessment tools and the length of time commitment it would take one to respond to the questionnaire which informed the data collection time points of day 1, first week and day 90. This strategy was to strike a balance between the feasibility of maximizing response rates while taking caution not to overload the already scare human resources of traffic law enforcement, trainees and clinicians at accident and emergency departments. Further engagements will be made during RTTDC training sessions and audit meetings during which bidirectional feedback on team performance, challenges with individual team dynamics and proposed areas of improvements to help shape future training.

### Data management {19}

Due to foreseen network breakdowns in rural centers, data will be collected in hard copies, coded and entered into the Research Electronic Data Capture (REDCap) secure virtual network hosted by the University of Turku. Data entry will be performed by outcome assessors who will be issued with confidential login password for REDCap. Later, the data will be exported to Stata version 15.0 for analysis. The REDCap software was preferred due its presumed secure web-based intuitive interface for validated data capture, offering an additional advantage to prohibit dual entry, restrict values, calibrate data ranges, and retrieve audit trails for tracking data manipulation [44]. Further, the software permits seamless data export and download procedures that are compatible with Stata, while allowing for data integration and interoperability with external sources [45]. Any data errors will be resolved during weekly audit meetings in reference to the online codebook with visible description of variables accessible to the principal investigator and all outcome assessors.

### Confidentiality {27}

To protect participants’ confidentiality before, during, and after the trial, all hard copy data collection items will be kept under lock and key and will bear unique non identifying codes. Soft copies will be kept in password protected computers with a second layer protection login password required for Redcap, only accessible by the study team. The final data sets will be anonymized prior to archive and publication. Hard copies of data will be destroyed six months after completion of the trial.

### Plans for collection, laboratory evaluation and storage of biological specimens for genetic or molecular analysis in this trial/future use {33}

There are no plans to collect any biological laboratory specimens either for purposes of storage, genetic or molecular analysis in this trial.

## Statistical methods

### Statistical methods for primary and secondary outcomes {20a}

#### Primary (process) outcomes

The difference in (i) mean prehospital, and (ii) referral-exit intervals (standard deviation) between intervention and control groups will be compared at 95% CI using a two sample t-test which is robust for normality deviations. The normality of distribution will be assessed using the Shapiro-Wilks test whereas equality of variance will be determined using Levene’s test. A difference in means of 60 minutes (1 hour) will be considered clinically meaningful in accordance with the golden hour principle [32].

#### Secondary (patient) outcomes

i. To compare morbidity of musculoskeletal injuries, the validated 10-item Trauma Expectation Factor Score (TEFS) at baseline and 10-item Trauma Outcome Measure Score (TOMS) at 90 days will be used [29]. The mean difference in TEFS and TOMS will be compared between groups using the two sample t-test at 95%. Further, the TOMS will be dichotomized into favorable (TOMS≥TEFS) and unfavorable (TOMS<TEFS) outcomes in accordance with previous studies [29]. The difference in these proportions between the intervention and control groups will be compared using adjusted Chi-square test if the expected counts are >5, otherwise by use of Fisher’s exact test at 95% CI. Further, we shall perform subgroup analyses to determine and control for factors that could be associated with unfavorable TOMS using individual level between-within mixed effects regression models which inherently overcomes the effect of smaller clusters and permits adjustment for covariates in a single stage, assuming equal allocation. The fixed effect variables will be the treatment arms (intervention vs. control) as the unit of analysis, with the odds ratios and their corresponding 95% CI as a direct estimate of the effect size. The covariates will include age, sex, education level, occupation, employment status, marital status, commute distance, road user category, injury severity score based on Kampala Trauma Score (KTS), presence or absence of fracture, nature of fracture if present, number of serious injuries, and treatment dichotomized surgical versus conservative.
ii. To compare morbidity of neurological injuries, the validated Glasgow Outcome Scale (GOS) at 90 days post injury will be used. The mean difference in GOS will be compared between groups using the two sample t-test. Further, the GOS will be dichotomized into favorable (GOS=4 or 5) and unfavorable (GOS=3 or 2 or 1) outcomes in accordance with previous studies [7]. The difference in proportions between the intervention and control group will be compared using adjusted Chi-square test if the expected counts are >5, otherwise by use of Fisher’s exact test at 95% CI.
iii. For all-cause mortality, the difference in proportions of all deaths at 90 days post-injury will be compared between intervention and control arms using Chi-square test if the expected counts are >5, otherwise by use of Fisher’s exact test at 95% CI. Lastly, we shall conduct subgroup analyses to determine and control for factors associated with all-cause mortality using individual and cluster level between-within mixed effects regression models. The covariates will include age, sex, injury mechanisms, mode of arrival, pre-hospital interval, and referral decision to hospital exit interval (dichotomized as ≤1hr or >1 hr.), pre-hospital first aid status, comorbidities, injury severity score based on Glasgow Coma Score (GCS) at baseline, multiplicity of injuries, Head and Brain CT based diagnosis and neurosurgical intervention. These variables have been found to influence outcomes following neurological trauma in previous studies [7]. All variables will be analyzed for confounding and effect modification using the Mantel-Haenszel statistics to probe necessary interaction terms. Variables with p-value of ≤0.2 at bivariate will be included in multivariable analysis. The median (IQR) of time to event i.e., from accident incident to death (in days) will be stratified by treatment arm and the difference compared using Wilcoxon two sample test. All analyses will be performed using Stata 15.0 and where appropriate, we shall use graphics such as box plots to visualize the data.

### Interim analyses {21b}

The Uganda National Council for Science and Technology accessed a preliminary report of interim analysis which were performed during 28 August 2022 when the half of the sample size (n=500) was attained and at its discretion recommended continuation of the study.

### Methods for additional analyses (e.g., subgroup analyses) {20b}

Subgroup analyses will be performed for: (i) varying road user categories (pedestrian, passenger, motorcyclists), (ii) injury mechanisms (motorcycle-motorcycle crash, motorcycle-pedestrian crash, motorcycle-static object, motorcycle-car crash), (iii) varying injury severities (mild, moderate, or severe based on the Kampala Trauma Score and Glasgow Comma Scale) and (iv) multiplicity of serious injuries (one or multiple). To determine the training-effect heterogeneity across time periods, we shall use an extension of Hussey and Hughes fixed effects model [46], to determine whether the intervention effect differs for each training period as summarized in the schematic representation of study design in Fig. 4.

**Fig. 4:**
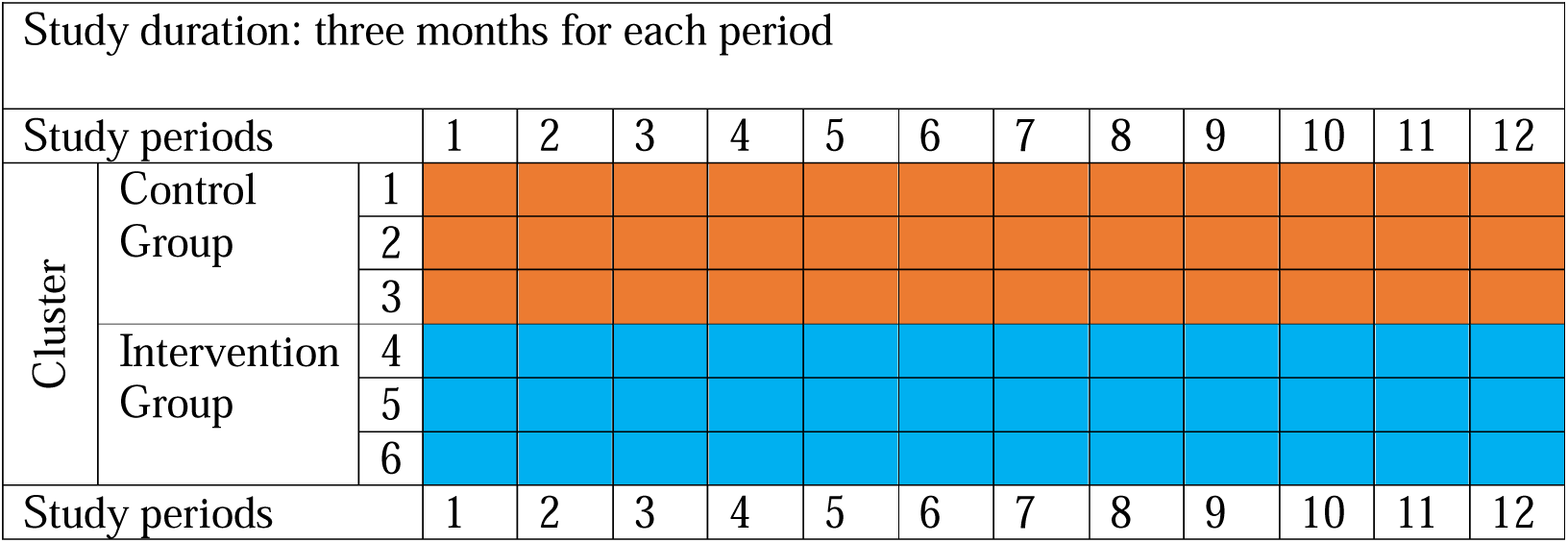
Schematic representation of the study design.

For ancillary study on provider outcomes, the difference in pre- and post-training mean scores will be computed at 95% CI if the data are normally distributed otherwise the difference in median and interquartile range will be reported. For the qualitative ancillary study on the perceived barriers to injury care, directed content analysis of themes of transcribed data will be collated manually and presented as percentages.

### Methods in analysis to handle protocol non-adherence and any statistical methods to handle missing data {20c}

We shall impute values for participants with missing end points such as lost to follow-up, withdrew consent, and crossovers. The baseline sociodemographic and clinical characteristics of such participants will be compared between the two treatment groups (intervention vs. control) and to those retained within the treatment groups. For individual participants, crossover from intervention to control group or vice versa due to any reason will result in ultimate discontinuation from the trial.

### Plans to give access to the full protocol, participant level-data and statistical code {31c}

The full protocol will be published open access and anonymized participant-level datasets, and statistical codes will be shared publicly through a permanent weblink that will be provided by the publishing journal. Since the primary country of recruitment which approved and registered the study prior recruitment lacked a publicly available electronic research register, this protocol was retrospectively registered with WHO approved Pan African Clinical Trial Registry (PACTR202308851460352).

## Oversight and monitoring

### Composition of the coordinating centre and trial steering committee {5d}

The principal investigator and a central study administrator will form a steering committee that will run the day-to-day activities of the trial. The trial will have two onsite overseers and two off-site supervisors. In addition, the principal investigator will provide organizational support to rural trauma teams at the intervention study centers through weekly audit meetings. Each of the rural trauma teams is composed of a road traffic law enforcement professional, third year medical student, fifth year medical student, intern doctor or nurse and a specialty surgery resident.

### Composition of the data monitoring committee, its role and reporting structure {21a}

The Research and Ethics Committee of Mbarara University of Science and Technology is the designated independent data monitoring committee (DMC) that will oversee this trial under reference number (MUREC 1/7; 05/5-19). The committee reports directly to the Uganda National Council for Science and Technology. The council at its discretion can recommend continuation, amendments, or termination of the trial at any time.

### Adverse event reporting and harms {22}

Immediate medical concerns from participants will be reported to their attending clinicians. Any reported adverse events and other unintended effects of trial interventions or trial conduct will be reported to the trial monitoring committee.

### Frequency and plans for auditing trial conduct {23}

The trial monitoring committees independently conduct annual and impromptu audits and may choose to extend or terminate the trial at any time. Such situations that could warranty termination include adverse events directly attributable to the study or the unlikely event of the trial achieving a meaningful difference in study outcomes between groups. The investigators will submit annual progress reports to the committees each year as part of continuing review.

### Plans for communicating important protocol amendments to relevant parties (e.g., trial participants, ethical committees) {25}

Any protocol amendments will be submitted to the data monitoring committee. Any approved amendments will be communicated to Uganda National Council for Science and Technology, trial participants, and updated in the registries within five working days following approval.

## Ancillary studies

This trial was approved with two ancillary studies. The first is to assess the effect of RRTDC on providers knowledge, and the second is a qualitative study that will assess the barriers to injury care as perceived by the traffic law enforcement and medical trainee frontliners and those encountered in real-time management of patient participants. The data collection team for the qualitative study will be surgery residents at the six regional referral hospitals who will be blinded by the cluster/treatment allocation. The results of the ancillary studies will be collected and analyzed separately and will be concealed from the trial team until the analysis of the main clinical trial findings is nearly complete. We hope that the findings of ancillary studies will complement and inform the interpretation of the trial findings.

### Dissemination plans {31a}

Participants for the RTTDC training will get feedback after the post-test questionnaire evaluation. Patient participants will be advised on the next plans of management during follow-up calls or through out-patient consultations. Before study findings will be presented in scientific conferences and published in peer reviewed journals, a copy of findings in the final bound report will be submitted to the participating hospital main libraries, departments of surgery and Internal Review Boards of Mbarara University of Science and Technology, Kampala International University and Uganda National Council for Science and Technology. The implications of findings will be shared with to authorities including interns and medical students’ associations, district health officers, hospital directors, regional traffic police commanders and chairpersons of the motorcycle riders’ associations in the respective regions, and with the rural trauma networks of first responders. Priority will be made to present the preliminary findings at an international conference hosted in Uganda. Anonymized participant datasets from this trial will be archived on a publicly accessible permanent weblink that will be provided by the publishing journal within 12 months from the actual date of completion of the trial.

## Discussion

This trial will provide a comparison of (i) the prehospital time interval from accident scene to hospital admission, (ii) the time interval from referral decision to hospital exit, (iii) all-cause mortality, and (iv) morbidity of patients with motorcycle-related orthopedic and neurological injuries presenting at RTTDC trained (intervention) versus standard care (control) study centers. The expected outcome of this trial is to reveal the overall impact of the rural trauma team development and training on the clinical process time efficiency and patient centered outcomes musculoskeletal and neurological injuries in controlled clinical settings, using appropriate injury severity scores, trauma outcome and process measures. The results of this trial could inform the design, implementation, and scalability of rural trauma team development in similar low-resource settings.

## Study strengths and limitations

To the best of authors knowledge, this is the first cluster randomized controlled trial using prospectively collected data to compare the effect of rural trauma team development course training and standard care on patient outcomes and process measures in LMICs. Whereas the authors are ambitious to assume that the study centers are homogeneous, the heterogeneity between the six participating regional referral hospitals could attract bias and affect study estimates. Further, the relatively smaller number of clusters could affect the power of the study. However, to randomize six out of the 17 trauma centers was within in ethical constraints as this sample already represents 35.3% of Uganda’s regional referral hospitals. To overcome the statistical power bias limitation, the sample size was inflated for individual randomization to cater for the design effects in accordance with Hooper et al [43]. Moreover, the authors plan to use mixed effects regression models which permit adjustment for covariates both at cluster and individual levels in a single stage approach, and making use of t-test that is robust to normality deviations. According to Borhan et al [47], the mixed effects models are deemed suitable for smaller clusters in situations where the primary outcomes are continuous as it is the case for this trial. Further, we hope that the ancillary studies with mixed methods will add value to the overall interpretation of the trial results, through raising the often unheard voices of sectors outside health such as the traffic law enforcement professionals.

## Trial status

The actual recruitment of the first patient participant began 01 September 2019. Last follow-up by 27 August 2023. Post-trail care including linkage to clinical, social support and referral services to be completed by 27 November 2023. This is protocol version V (2023).

## Supporting information

SPIRIT Checklist

Ethical Approval

## Data Availability

All data produced in the present work are contained in the manuscript

https://pactr.samrc.ac.za/TrialDisplay.aspx?TrialID=25763

## Abbreviations

RTTDC: Rural Trauma Team Development Course
ATLS: Advanced Trauma Life Support
PTC: Primary Trauma Course
ETC: European Trauma Course
LMICs: Low-and Middle-Income Countries
MUST: Mbarara University of Science and Technology
UNCST: Uganda National Council for Science and Technology
GCS: Glasgow Coma Scale
GOS: Glasgow Outcome Scale
TEFS: Trauma Expectation Factor Score
TOMS: Trauma Outcome Measure Score
KTS: Kampala Trauma Score

## Acknowledgements

The authors are grateful to the staff of the research and ethics committee of Mbara University of Science and Technology for their technical and ethical guidance during the early phases of this project design.

## Declarations

### Authors’ contributions {31b}

HL: Principal investigator, Conceptualization, Data curation, Investigation, Methodology, Project administration, Resources, Wring-original draft; HL, MM: Formal analysis, Software, Visualization; RS, PK, TB, JPP, MLW: Validation, Writing-review, and editing; TB, JPP, MLW: Supervision, Funding acquisition. All authors read and approved the final manuscript. In addition, we plan to outsource a blinded external biostatistician to validate the final data analyses and will be included as a co-author on the resulting manuscripts. All authorship will be in accordance with the International Committee of Medical Journal Editors guidelines.

### Availability of data and materials {29}

All contents of the SPIRIT checklist are contained within this protocol. The anonymized datasets arising from this study will be archived and made publicly available through a permanent weblink to a repository that will be provided by the publishing peer reviewed journal.

### Ethics approval and consent to participate {24}

This study was approved and registered by the research and ethical committee of Uganda National Council for Science and Technology (Ref. No. SS 5082) prior recruitment. Written informed consent will be obtained from all study participants.

### Consent for publication {32}

All participants or their legally authorized representatives will endorse a predesignated consent form with their signatures prior recruitment. Model consent forms for trainee and patient participants are available as online supplementary material 1 and 2 respectively.

## Authors’ information

HL: Chief of Surgery, Kiryandongo Hospital, Kigumba, Uganda; Doctoral Researcher, Injury Epidemiology and Prevention Research Group, Turku Brain Injury Center, Department of Clinical Neurosciences, Turku University Hospital and University of Turku, Turku, Finland.

MM: Biostatistician and Applied Epidemiologist, Injury Epidemiology and Prevention (IEP) Research Group, Turku Brain Injury Centre, Department of Clinical Neurosciences, Turku University Hospital and University of Turku, Turku, Finland.

RS: Associate professor of Surgery; Director of Medical Education, Training and Research, Department of Surgery, Mengo Hospital, Kampala, Uganda; Chair of Ethics and Disciplinary Committee, Uganda Medical and Dental Practitioners’ Council.

PK: Professor of Surgery; Fellow of High Education Academy, and Vice Chancellor of Mother Kevin Postgraduate Medical School, Uganda Martyr’s University, Nkozi, Uganda.

TB: Alexander von Humboldt University Professor of Global Health and Director Heidelberg Institute of Global Health, the University of Heidelberg, Germany; Adjunct professor of Global and Population Health at Harvard T.H. CHAN School of Public Health (USA) and Senior Faculty-Africa Health Research Institute (ARHI), South Africa.

JPP: Adjunct Professor of Neurosurgery; Head of Department of Neurosurgery, Brain Injury and Rehabilitation, Turku University Hospital and University of Turku, Turku, Finland; Adjunct Professor of Neurotraumatology, University of Helsinki, Helsinki, Finland.

MLW: Adjunct Professor and Head of Injury Epidemiology and Prevention, Heidelberg Institute of Global Health (HIGH), University Hospital and University of Heidelberg, Heidelberg, Germany.

