## Supplemental Material 1 for "Effect of rural trauma team development on outcomes of motorcycle related injuries: A protocol for a multi-center cluster randomized controlled clinical trial (The MOTOR trial)"

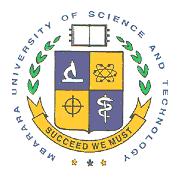
**MBARARA UNIVERSITY OF SCIENCE AND TECHNOLOGY**

**RESEARCH ETHICS COMMITTEE**

INFORMED CONSENT FORM (TRAINEE PARTICIPANTS)

This document outlines the research study and expectations for potential participants. It should be written in layman terms and typed on MUST-REC letterhead.

**Instructions**

1. The wording of this document should be directed to the potential participant not MUST-REC.
2. If a technical term must be used, then define it the first time it is used and any acronyms or abbreviations used should be spelled out the first time they are used.
3. All the sections of this document must be completed without any editing or deletions.
4. Please use a typing font that is easily distinguishable from the questions of this form. Preferably the font size should be 12.

**Study title** – This should be the same as on all other documents related to the study.

**Lay title:** The use of allied health and law enforcement trauma team registries: Lessons from a novel motorcycle trauma registry and surveillance system in Uganda

**Scientific title:** An examination of the impact of rural trauma team development and training on the outcomes of motorcycle related orthopedic and neurological injuries: A protocol for a multi-center cluster randomized controlled clinical trial (The MOTOR trial).

**Principal Investigator(s)**

Lule Herman

**Introduction**

What you should know about this study:

1. You are being asked to join a research study.
2. This consent form explains the research study and your part in the study.
3. Please read it carefully and take as much time as you need.
4. You are a volunteer. You can choose not to take part and if you join, you may quit at any time. There will be no penalty if you decide to quit the study.

**Brief background to the study**

Limb and head injuries are a burden to passengers, motorcycle riders and people who walk on streets. The increasing number of motorcycle-related accidents could be a contributing factor. This study looks at the commonest injuries resulting from such accidents, the severity and outcome of such injuries and if forming a trained team at parish level can help in improving the quality of information obtained in treating such injuries and the overall outcome. The study seeks to estimate the burden of motorcycle related accidents in our local context, contributing factors to poor outcome and contribute to management of such patients through training response teams.

**Purpose of the research project**

Include a statement that the study involves research, estimated number of participants, an explanation of the purpose(s) of the research procedure and the expected duration of the subject's participation.

This project is part of the requirements for an academic award and is expected to last for four years (2019 to 2023). In total about 500 trainee participants are targeted, meaning about 166 at each of the 3 participating hospitals in a span of 4 interviews (trainings) at 3 months interval corresponding to internship rotations. Of the 500 trainee participants, 66 (13.2%) will be road traffic police officers, 30 (6.0%) intern doctors, 140 (28.0%) fifth year and 264 (52.8%).

**Why you are being asked to participate?**

Explain why you have selected the individual to participate in the study.

You have been selected because this study is targeting people who are involved in care and response to patients involved in road traffic crashes such as motorcycle related accidents. If you are a traffic police officer, medical student in surgery clinical rotation, intern doctor/nurse or allied health professional you qualify for inclusion for this training.

**Procedures**

Provide a description of the procedures to be followed and identification of any procedures that are experimental, clinical etc. If there is need for storage of biological (body) specimens, explain why, and include a statement requesting for consent to store the specimens and state the duration of storage.

As a participant, you will benefit from the training since you are part of the response team (police, medical student, intern nurse or doctor). You will be given a brief overview of the study and why forming a response team might be important in improving injury data and treatment outcome of such patients. If you decide to participate, you will be asked to provide information about your cadre and contact details (phone number and email address). You will then be asked to sign a consent form document and later be enrolled for a pre-test questionnaire. You will undergo rural trauma team development training during which you will be assigned to a team and subsequently complete post training follow-up questionnaire.

You will also be given a chance to tell us about the main barriers to injury care at your hospital of attachment and provide feedback on how you feel about this training. You might be contacted by a member of our team or your team member in future to discuss team challenges.

**Risks or discomforts**

Describe any reasonably foreseeable risks or discomforts-physical, psychological, social, legal or other associated with the procedure, and include information about their likelihood and seriousness. Discuss the procedures for protecting against or minimizing any potential risks to the subject. Discuss the risks in relation to the anticipated benefits to the subjects and to society.

Apart from the discomfort that might arise from recalling the bad experience of events that could have occurred during an accident in which you or your relatives were involved, there are no additional health risks posed by this study/training since it only requires you to respond to questions and be available for future contact.

**Benefits**

Describe any benefits to the subject or other benefits that may reasonably be expected from the research. If the subject is not likely to benefit personally from the experimental protocol note this in the statement of benefits.

As a participant, you will access the most recent information about the burden attributed to motorcycle related road traffic accidents and contribute to the existing body of knowledge in this field. Besides, you will undergo simulated training that could improve your skills and earn a certificate of participation. In the end, you will benefit when you become a member of trauma team network and when recommendations based on this study findings are implemented by policy makers, regarding prevention and improvement of case management of motorcycle injured patients through emergency response teams.

**Incentives or rewards for participating**

It is assumed that there are no costs to subjects enrolled in research protocols. Any payments to be made to the subject, e.g., travel expenses, token of appreciation for time spent, must also be stated, including when the payment will be made.

As a participant, you need to understand that your participation in this study is voluntary and that there is no economic gain from this study. Only a transport refund based on your commute distance will be reimbursed for your convenience.

**Protecting data confidentiality**

Provide a statement describing the extent, if any, to which confidentiality or records identifying the subjects will be maintained. If data is in form of tape recordings, photographs, movies or videotapes, researcher should describe period of time they will be retained before destruction. Showing or playing of such data must be disclosed, including instructional purposes.

The investigator will protect all the information obtained with a password and lockable shelves with a key only known to him and will not reveal any of your personal information without your consent. Hard copies of information collected will be destroyed at the end of study period whereas anonymous soft copy will be archived.

**Protecting subject privacy during data collection**

Describe how the privacy of the participant will be ensured during the process of data collection.

Your name will never appear anywhere on any documents that will arise from this study, instead a unique number will be assigned to you. Only investigators will have access to this information

**Right to refuse or withdraw**

Include a statement that participation is voluntary and that refusal to participate will involve no penalty or loss of benefits to which the subject is otherwise entitled.

Since your participation is voluntary, nothing will happen if you refuse to participate in this study; or if you withdraw from the study.

**What happens if you leave the study?**

Include a statement that the subject may discontinue participation at any time without penalty or loss of benefits.

You will not incur any loss of benefits to which you would otherwise be entitled. You are free to withdraw from the study at any time without explaining your actions.

**Who do I ask/call if I have questions or a problem?**

Include contact for the researcher and Chairperson, MUST-REC.

If you have any queries/problem at any time about this study, contact Dr. Lule Herman C/o Turku Brain Injury Centre, Division of Clinical Neurosciences, Faculty of Medicine, University of Turku or reach him on Tel.0775656222/0758997877 or on E-mail address

OR Contact Prof. Ssebuufu Robinson, Dean and Executive Director, Kampala International University-Western Campus P.o.Box 71-Ishaka on Tel: 0772507248 or

OR Contact Dr. Francis Bajunirwe, Chairman Mbarara University of Science and Technology (MUST) IRC P.O.Box 1410 Mbarara Tel: 0485433795

**What does your signature or thumbprint on this consent form mean?**

Your signature on this form means

- You have been informed about this study’s purpose, procedures, possible benefits and risks
- You have been given the chance to ask questions before you sign
- You have voluntarily agreed to be in this study

_ _ _ _ _ _ _ _ _ _ _ _ _ _ _ _ _ _	_ _ _ _ _ _ _ _ _ _ _ _ _ _ _ _ _ _	_ _ _ _ _ _ _ _ _ _ _ _

Name of adult participant	Signature of participant or	Date

Legally authorized representative

_ _ _ _ _ _ _ _ _ _ _ _ _ _ _ _ _ _	_ _ _ _ _ _ _ _ _ _ _ _ _ _ _ _ _ _	_ _ _ _ _ _ _ _ _ _ _ _

Name of person obtaining	consent	Signature	Date

_ _ _ _ _ _ _ _ _ _ _ _ _ _ _ _ _ _	_ _ _ _ _ _ _ _ _ _ _ _ _ _ _ _ _ _	_ _ _ _ _ _ _ _ _ _ _ _

Print Name of witness	Signature or thumbprint or mark	Date
