## Supplemental Material 2 for "Effect of rural trauma team development on outcomes of motorcycle related injuries: A protocol for a multi-center cluster randomized controlled clinical trial (The MOTOR trial)"

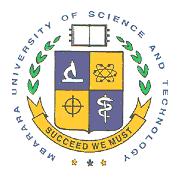
**MBARARA UNIVERSITY OF SCIENCE AND TECHNOLOGY**

**RESEARCH ETHICS COMMITTEE**

INFORMED CONSENT FORM (PATIENT PARTICIPANTS)

This document outlines the research study and expectations for potential participants. It should be written in layman terms and typed on MUST-REC letterhead.

**Instructions**

1. The wording of this document should be directed to the potential participant not MUST-REC.
2. If a technical term must be used, then define it the first time it is used, and any acronyms or abbreviations used should be spelled out the first time they are used.
3. All the sections of this document must be completed without any editing or deletions.
4. Please use a typing font that is easily distinguishable from the questions in this form. Preferably the font size should be 12.

**Study title** – This should be the same as on all other documents related to the study.

**Lay title:** The use of allied health and law enforcement trauma team registries: Lessons from a novel motorcycle trauma registry and surveillance system in Uganda

**Scientific title:** An examination of the impact of rural trauma team development and training on the outcomes of motorcycle related orthopedic and neurological injuries: A protocol for a multi-center cluster randomized controlled clinical trial (The MOTOR trial)

**Principal Investigator(s)**

Lule Herman

**Introduction**

What you should know about this study:

1. You are being asked to join a research study.
2. This consent form explains the research study and your part in the study.
3. Please read it carefully and take as much time as you need.
4. You are a volunteer. You can choose not to take part and if you join, you may quit at any time. There will be no penalty if you decide to quit the study.

**Brief background to the study**

Limb and head injuries are a burden to passengers, motorcycle riders and people who walk on streets. The increasing number of motorcycle-related accidents could be a contributing factor. This study looks at the commonest injuries resulting from such accidents, the severity and outcome of such injuries and if forming teams of people who encounter such patients first especially in remote areas can help in improving the quality of information obtained, care for such injuries and their overall outcome. The study seeks to estimate the burden of motorcycle related accidents in our local context, contributing factors to poor outcome and contributing to management of such patients in this aspect.

**Purpose of the research project**

Include a statement that the study involves research, estimated number of participants, an explanation of the purpose(s) of the research procedure and the expected duration of the subject's participation.

This project is part of the requirements for an academic award and is expected to last for four years tentatively (2019 to 2023). We hope to recruit one to two patient participants per day in each of the participating hospitals. Each participant will be followed-up for a period of 3 months to assess how they are recovering and coping with their injuries. We shall also note any challenges your doctors might face in giving you treatment.

**Why you are being asked to participate?**

Explain why you have selected the individual to participate in the study.

You have been selected because this study is targeting patients involved in motorcycle-related accidents. If you have any injuries as result of a motor cyclist knocking another motorcycle, motor cyclist knocked by a car, you were walking on streets and knocked by a motorcycle or passenger on a motorcycle being involved in road traffic accident, you qualify for inclusion.

**Procedures**

Provide a description of the procedures to be followed and identification of any procedures that are experimental, clinical etc. If there is need for storage of biological (body) specimens, explain why, and include a statement requesting for consent to store the specimens and state the duration of storage.

If you are participating as a patient, your breathing, blood pressure, level of consciousness and serious injuries will be assessed.

You will be given emergency treatment like stopping the bleeding and pain killers based on your injuries until the doctor thinks you are stable before you can be enrolled into the study.

You will then be enrolled, asked a few questions regarding your age, profession, level of education, when and where the accident took place, how you arrived at the hospital and any care you received before hospital.

You will also be given a chance to tell us how you feel these injuries are going to affect your life and work today and 3 months from now. You will therefore be asked to provide a contact (for you and

Your attendant) for a curtsey phone call to find out how you are doing or remind you to come for your appointment and link or referral to any assistance you might need thereafter to recover from your injuries.

Once the doctor suspects you have a broken leg, or injured your head, you will be sent to “X-ray” or scan to confirm. This service is usually available freely in government facilities. The investigator will only meet this cost if the government free “X-ray” services are not available/functional. When you are ready to receive the results, your Drs will tell you the results and the next plan of management which the researcher will record on the questionnaire. You will however retain your “X-ray” film.

**Risks or discomforts**

Describe any reasonably foreseeable risks or discomforts-physical, psychological, social, legal, or other associated with the procedure, and include information about their likelihood and seriousness. Discuss the procedures for protecting against or minimizing any potential risks to the subject. Discuss the risks in relation to the anticipated benefits to the subjects and to society.

Apart from the discomfort that might arise from recalling the bad experience of events that occurred during the accident, there are no additional health risks posed by the study since it only requires you to respond to questions, undergo medical examination to detect any life threatening injuries that needs urgent attention and “X-ray” assessment (only if necessary) in accordance with trauma management guidelines to ascertain and confirm the nature of your broken leg or head damage.

**Benefits**

Describe any benefits to the subject or other benefits that may reasonably be expected from the research. If the subject is not likely to benefit personally from the experimental protocol note this in the statement of benefits.

As a participant, you will be linked to appropriate care of your injuries, access most recent information about the burden attributed to motorcycle related road traffic accidents and contribute to the existing body of knowledge in this field besides better understanding of the nature of your injuries. In the end, you will benefit when recommendations based on this study findings are implemented by policy makers, regarding prevention and improvement of case management of motorcycle injured patients

**Incentives or rewards for participating**

It is assumed that there are no costs to subjects enrolled in research protocols. Any payments to be made to the subject, e.g., travel expenses, token of appreciation for time spent, must also be stated, including when the payment will be made.

As a participant, you need to understand that your participation in this study is voluntary and that there is no economic gain from this study. You will get ten thousand shillings as compensation for your time and transport refund if your follow-up study visits are not related to routine patient care.

_ _ _ _ _ _ _ _ _ _ _ _ _ _ _ _ _ _	_ _ _ _ _ _ _ _ _ _ _ _ _ _ _ _ _ _	_ _ _ _ _ _ _ _ _ _ _ _

Name of adult participant	Signature of participant or	Date

Legally authorized representative

_ _ _ _ _ _ _ _ _ _ _ _ _ _ _ _ _ _	_ _ _ _ _ _ _ _ _ _ _ _ _ _ _ _ _ _	_ _ _ _ _ _ _ _ _ _ _ _

Name of person obtaining	consent	Signature	Date

_ _ _ _ _ _ _ _ _ _ _ _ _ _ _ _ _ _	_ _ _ _ _ _ _ _ _ _ _ _ _ _ _ _ _ _	_ _ _ _ _ _ _ _ _ _ _ _

Print Name of witness	Signature or thumbprint or mark	Date
