## Supplemental Material 3 for "Effect of rural trauma team development on outcomes of motorcycle related injuries: A protocol for a multi-center cluster randomized controlled clinical trial (The MOTOR trial)"

Supplementary Material 3: Process measures and participant outcomes

| Category | Type | Outcome | Specific Measurement Variable | Analysis Metric | Method of Aggregation | Time Point |
| --- | --- | --- | --- | --- | --- | --- |
| Process Measures | Primary | Crash scene-admission interval. | Time taken in hours from crash scene to admission as documented on police crime scene records, CCTV cameras, and emergency department admission records. | Final Value | Mean/Median/Difference in means (SD) between intervention and control group. | Day 1of admission. |
|  |  | Referral Exit-Interval | Time taken in hours from decision to refer until the patient exits hospital as extracted from case files, referral forms and handover reports from ambulance drivers and accompanying nurses. | Final Value | Mean/Median/ Difference in means (SD) or median (IQR) between intervention and control group. | Day 1 of admission until referral point. |
| Patient Participant Outcomes | Secondary | All-cause mortality within 90-days from the time of injury. | Death as reported in case files, death certificates, post-mortem reports, or mortuary records. | Final value | Proportions/Difference in proportions between intervention and control groups. | 90-days from the time of injury. |
|  | Secondary | Time to all-cause mortality during follow-up period. | Time in days to death as documented by the treating physician in case files, death certificates, mortuary records. | Final value | Median/Difference in median (IQR), Survival analysis, hazard | At the end of follow-up. |
|  | Secondary | Cause specific in-hospital mortality for neurological injuries within 90 days from time of injury. | Categorical cause of death as documented in case files, death certificates, post-mortem reports, or mortuary records by a certified physician. | Final value | Proportions/Difference in proportions between intervention and control group. | At the end of follow-up. |
|  | Secondary | Morbidity of musculoskeletal injuries | Level of functional and physical disability as reported by the patients based on trauma expectation factor score (TEFS) at admission and trauma outcome measure score (TOMS) at 90 days as reported by patients during physical interviews, home visits and telephonic follow-up. | Final value | Mean/Median/ Difference in means (SD) or median (IQR) between intervention and control group. | (TEFS) during first week of admission and (TOMS) at  90-days post injury. |
|  | Secondary | Cause-specific unfavorable trauma outcome measure for musculoskeletal injuries. | Secondary analyses after case definition of favorable trauma outcome measure as (TOMS) at 90 days ≥ (TEFS) at first week of admission. | Final value | Proportions/ Difference in proportions between intervention and control group. | At 90 days post injury. |
|  | Secondary | Morbidity of neurological injuries. | Level of neurological disability based on admission Glasgow Coma Scale (GCS) and Glasgow outcome scale (GOS) at 90 as assessed and documented by the attending physicians during physical follow-up days. | Final value | Mean/Median/ Difference in means (SD) or median (IQR) between intervention and control group. | GCS at time of admission and GOS at 90 days post injury. |
| Provider (Trainee) Participant Outcomes | Tertiary  (ancillary) | Trauma knowledge gain/retention. | Effect of training on medical trainees’ knowledge retention as assessed based on pre -and post-training trauma related MCQs | Final values | Mean/Median/Difference in mean (SD) or median (IQR) MCQ percentage scores before and after the training. | Pre-test MCQs First day of the training and at Post-test MCQs at 90 days after the training. |
|  | Tertiary  (ancillary) | Barriers to Injury Care | Barriers to injury care as reported by trainee care providers | Final values | Proportions/Difference in proportions of categorical themes between intervention and control group | Throughout the course of follow-up |
