## Supplemental Material 4 for "Effect of rural trauma team development on outcomes of motorcycle related injuries: A protocol for a multi-center cluster randomized controlled clinical trial (The MOTOR trial)"

### Form 1

Please complete the survey below.

Thank you!

| | |
|---|---|
| Date of admission | ________________________________<br>(Indicate the date of admission eg first visit (Date-Month-Year) |
| Date of follow-up | ________________________________<br>(Indicate date of follow-up at 12 weeks) |
| Hospital facility | ◯ Kiryandongo Regional Referral Hospital<br>◯ Jinja Regional Referral Hospital<br>◯ Hoima Regional Referral Hospital<br>◯ Mubende Regional Referral Hospital<br>◯ Fortportal Regional Referral Hospital<br>◯ Kampala International University Teaching Hospital<br>(Hospital facility where patient was attended to) |
| Hospital Category | ◯ (intervention group)<br>◯ Control group<br>(Assign hospital as case control) |
| Age (Years) | ________________________________<br>(Age in years) |
| Informant | ◯ Patient<br>◯ Regally Authorized Representative (Guardian/Parent)<br>(Indicate who provided the information eg Parent for under age (< 18 years) or mentally incapacitated patients) |
| Sex | ◯ Male<br>◯ Female<br>(Sex) |
| Religious affiliation | ◯ Christian<br>◯ Moslem<br>◯ Other<br>(Enter religious affiliation if applicable) |
| Education Level | ◯ Primary education<br>◯ Secondary education (high school)<br>◯ Tertiary education (university)<br>(Enter level of education) |
| Marital status | ◯ Single<br>◯ Married<br>◯ Divorced<br>◯ other<br>(Marital status) |

| | |
|---|---|
| Occupation | ◯ Commercial motorcyclist (bodaboda)<br>◯ Peasant farmer<br>◯ Business man/woman<br>◯ Student<br>◯ Teacher<br>◯ Politician<br>◯ Priest/Pastor<br>◯ Engineer<br>◯ Taxi Driver<br>◯ Other<br>(what do you do for a living?) |
| Employment status | ◯ Formal paid employment<br>◯ Self-employed<br>◯ Unemployed<br>◯ Student<br>◯ Other<br>(Employment status) |
| History of alcohol consumption | ◯ Yes<br>◯ No<br>(Was there history of alcohol consumption prior to accident incident?) |
| Referral Status | ◯ Referred from a lower facility<br>◯ Self-referral<br>(How did the patient chose this health facility?) |
| Estimated commute distance (Km) from accident scene to facility | __________________________________<br>(Roughly how many kilometers from accident scene to this health facility) |
| Road user category | ◯ Passenger<br>◯ Pedestrian<br>◯ Motorcyclist<br>◯ Driver<br>(The capacity in which the participant was using the road at the time of accident incident) |
| Mechanism of injury | ◯ Motorcycle-motorcycle crash<br>◯ Motorcycle-pedestrian crash<br>◯ Motorcycle-car crash<br>◯ Motorcycle-static object crash<br>(Nature of crash) |
| Mode of arrival | ◯ By ambulance<br>◯ By taxi/motorcycle<br>◯ Other<br>(How did the patient arrive at the hospital?) |
| Estimated time lag from accident to arrival at hospital (hours) | __________________________________<br>(How many hours did it take you to reach the hospital?) |

| | |
|---|---|
| Pre-hospital care (first aid before arrival)? | ◯ Yes<br>◯ No<br>(Did the patient receive any first aid prior arrival?) |
| If yes, Who administered the first aid? | ◯ Health worker<br>◯ Lay-bystander/police<br>(Indicate who administered the first aid) |
| Evidence of chronic medical illness? | ◯ Yes<br>◯ No<br>(Does the participant has any medical illness?) |
| If yes, specify medical illness | ◯ Diabetes mellitus<br>◯ Hypertension<br>◯ HIV/AIDS<br>◯ Ischaemic heart disease<br>◯ Congestive heart failure<br>◯ Asthma<br>◯ Chronic obstructive airway disease<br>◯ Epilepsy<br>◯ COVID-19<br>◯ Stroke<br>◯ Malnutrition<br>◯ Tuberculosis<br>(Specify nature of medical illness if present) |
| Body system injured | ☐ Head<br>☐ Neck<br>☐ Chest<br>☐ Abdomen<br>☐ Pelvis<br>☐ Musculoskeletal<br>☐ Other<br>(Specify the body part that was injured (tick all applicable)) |
| Limb fracture present? | ◯ Yes<br>◯ No<br>(Indicate if a limb fracture is present) |
| If fracture is present, specify which bone (tick all that apply) | ☐ Tibia<br>☐ Femur<br>☐ Humerus<br>☐ Radius/Ulnar<br>☐ Other eg clavicle<br>(If the patient sustained any fractures, which bones were affected?) |
| Nature of tibial fracture (if present) | ◯ Open<br>◯ Closed<br>(Classify the fracture as open or closed if present) |
| Pattern of tibial fracture | ◯ Proximal 1/3<br>◯ Mid 1/3<br>◯ Distal 1/3<br>(Indicate which segment of tibial bone that fractured) |

| | |
|---|---|
| Is head injury present /suspected? | ◯ Yes<br>◯ No<br>(Indicate if head injury is present) |
| Signs and symptoms for suspecting head injury (Tick all that apply) | ☐ No helmet use<br>☐ High impact injury<br>☐ Loss of consciousness<br>☐ Post traumatic convulsions<br>☐ Post traumatic amnesia<br>☐ Post traumatic headache<br>☐ Alcohol or drug intoxication<br>☐ Projectile vomiting<br>☐ Visible injuries above clavicles<br>☐ CSF rhinorrhea / Otorrhea<br>☐ Focal neurological deficits<br>(Why is head injury suspected?) |
| Head and brain CT diagnosis? | ☐ Epidural Hematoma<br>☐ Subdural Hematoma<br>☐ Subarachnoid Hematoma<br>☐ Intraventricular Hemorrhage<br>☐ Intraparenchymal Hemorrhage<br>☐ Diffuse Axonal/Vascular Injury<br>☐ Cortical Contusions<br>☐ Skull fracture<br>☐ Normal CT results<br>☐ Other lesions e.g., maxillofacial<br>☐ CT results not conclusive<br>(If head and brain CT scan was obtained, indicate primary radiological diagnosis.) |
| Neurosurgical intervention | ☐ Craniotomy and clot evacuation<br>☐ Decompressive craniectomy<br>☐ Watchful waiting<br>(Indicate type of Neurosurgical intervention if known) |
| Total Glasgow coma score | ________________________________<br>(Indicate total GCS if head injury is present) |
| Glasgow coma scale (GCS) category | ◯ Mild Head Injury (GCS 13-15)<br>◯ Moderate Head Injury (GCS 9-12)<br>◯ Severe Head Injury (GCS 8 or less)<br>(Indicate severity of head injury (if applicable) based on GCS) |

| | No response | To pain | To verbal stimuli | Spontaneous |
|---|---|---|---|---|
| Eye opening | ◯ | ◯ | ◯ | ◯ |

| | No response | Incomprehensibl e speech | Inappropriate words | Confused conversation | Oriented |
|---|---|---|---|---|---|
| Verbal | ◯ | ◯ | ◯ | ◯ | ◯ |

| | No response | Extension in response to pain | Flexion in response to pain | Withdraws from pain | Localizes pain | Obeys command |
|---|---|---|---|---|---|---|
| Motor response | ◯ | ◯ | ◯ | ◯ | ◯ | ◯ |

Kampala Trauma Score (KTS) II

________________________________
(Indicate severity of other injuries by quoting the KTS)

Category of KTS

◯ Mild (9-10) ◯ Moderate (7-8)
◯ Severe (6 or less)
(Indicate injury severity based on KTS)

**KTS (A)**

| | < 5>55 | 5-55 |
|---|---|---|
| Age (years) | ◯ | ◯ |

**KTS (B)**

| | less or equal to 9 | equal or greater than 30 | 10-29 |
|---|---|---|---|
| Respiratory rate at admission (breaths per minute) | ◯ | ◯ | ◯ |

**KTS (C)**

| | Less or equal to 49 | 50-89 | > 89 |
|---|---|---|---|
| Systolic BP at admission (mmHg) | ◯ | ◯ | ◯ |

**KTS (D)**

| | Unresponsive (U) | Responds to Pain (P) | Responds to voice (V) | Alert (A) |
|---|---|---|---|---|
| Neurological Status | ◯ | ◯ | ◯ | ◯ |

**KTS (E)**

| | >one | One | None |
|---|---|---|---|
| Score for serious injuries | ◯ | ◯ | ◯ |

Decision (intention) on mode of treatment for the orthopedic and musculoskeletal injuries

◯ Conservative (watchful waiting, physical therapy, casting, braces, splints, pain medication)
◯ Operative (Open reduction and internal or external fixation, soft tissue repair requiring theatre)
(Indicate treatment modality)

Any barrier to receiving definitive injury care?

◯ Yes
◯ No

Does severity or multiplicity of injuries exceed local resources and capacity requiring referral?

◯ Yes
◯ No

Interval between referral decision and hospital exit (Hours)

________________________________

Barrier to receiving definitive care

- ☐ No barrier encountered in execution of definitive injury care
- ☐ Team barriers (leading to delays in emergency skilled team activation, identification, prioritization, recognition, timely referral of life threatening injuries)
- ☐ Individual barriers (leading to delays in decision to operate, initiate treatment, referral of injuries exceeding local capacity)
- ☐ In-hospital system barriers (leading to delays in securing necessary supplies eg oxygen, anesthetics, sutures, blood products, functional diagnostics, theatre space, intensive and critical care services)

(Specify any barrier to receiving definitive injury care encountered or documented for this particular patient)

#### Trauma Expectation Factor Score (TEFS) at admission

| | 0% | 25% | 50% | 75% | 100% |
|---|---|---|---|---|---|
| 12 weeks after surgery/treatment ,how painful do you expect your injury to be? | ○ | ○ | ○ | ○ | ○ |
| 12 weeks after surgery/treatment, how much do you expect your injury to interfere with your usual activity including prolonged standing, walking, climbing stairs, car driving, sleeping? | ○ | ○ | ○ | ○ | ○ |
| 12 weeks after surgery/treatment, how much do you expect your injury to interfere with your usual physical activity including work, house work, school, recreation/sports? | ○ | ○ | ○ | ○ | ○ |
| 12 weeks after surgery/treatment, how much do you expect your injuries to interfere with your usual activities of daily living, including eating, dressing, wearing shoes? | ○ | ○ | ○ | ○ | ○ |

| | | | | | |
|---|---|---|---|---|---|
| 12 weeks after surgery/treatment, how much do you expect your injury to interfere with your usual relationships, including family, friends and coworkers? | ◯ | ◯ | ◯ | ◯ | ◯ |
| 12 weeks after surgery/treatment, how much do you expect to cutdown on physical activities that are necessary to do including work, house work and school? | ◯ | ◯ | ◯ | ◯ | ◯ |
| 12 weeks after surgery/treatment, how much do you expect to cut down on the optional physical activity you enjoy doing, including sports, recreation, gardening? | ◯ | ◯ | ◯ | ◯ | ◯ |
| 12 weeks after surgery/treatment, how satisfied do you expect to be with your level of pain, physical function and disability? | ◯ | ◯ | ◯ | ◯ | ◯ |
| 12 weeks after surgery/treatment, how satisfied do you expect to be with 12 weeks after surgery/treatment, how satisfied do you expect to be with your physical appearance due to injuries you sustained? | ◯ | ◯ | ◯ | ◯ | ◯ |
| 12 weeks after surgery/treatment, how satisfied do you expect to be with your overall wellbeing? | ◯ | ◯ | ◯ | ◯ | ◯ |

Total Trauma Expectation Factor Score (TEFS) score

__________________________________
(Indicate the total trauma expectation factor at zero weeks score prior surgical care)

Injury outcome at 12 weeks

◯ Survived
◯ Died
◯ Lost to follow up
(what was the fate of the patient at 12 weeks?)

Day of death

__________________________________
(If died, indicate the death day from crash incident)

#### TOMS_12WEEKS

| | 0% | 25% | 50% | 75% | 100% |
|---|---|---|---|---|---|
| How painful is your injury today? | ◯ | ◯ | ◯ | ◯ | ◯ |
| How much does your injury currently interfere with your usual necessary activity eg prolonged standing, walking stairs, car driving and sleeping | ◯ | ◯ | ◯ | ◯ | ◯ |
| How much does your injury currently interfere with your usual physical eg work, house work school, recreation, sports ? | ◯ | ◯ | ◯ | ◯ | ◯ |
| How much does your injury currently interfere with your usual activities of daily living eg eating, dressing, putting on shoes? | ◯ | ◯ | ◯ | ◯ | ◯ |
| How much does your injury currently interfere with your usual relations eg family, friends, coworkers | ◯ | ◯ | ◯ | ◯ | ◯ |
| How much do you currently cut down necessary physical activities eg work, house work , school? | ◯ | ◯ | ◯ | ◯ | ◯ |
| How much do you currently cut down on physical activities you enjoy doing eg sports, recreation and gardening | ◯ | ◯ | ◯ | ◯ | ◯ |
| How satisfied are you with your current level of pain, physical function and disability | ◯ | ◯ | ◯ | ◯ | ◯ |
| How satisfied are you with the current appearance of your injury | ◯ | ◯ | ◯ | ◯ | ◯ |
| How satisfied are you with your current overall wellbeing | ◯ | ◯ | ◯ | ◯ | ◯ |

Total Trauma Outcome Measure Score (TOMS)

__________________________________
(Indicate the total trauma outcome measure score at 12 weeks post injury treatment)

Trauma outcome measure category

◯ Favorable trauma outcome measure (Total TOMS is Equal or Greater than TEFS)
◯ Unfavorable trauma outcome measure (Total TOMS is less than TEFS)
(Indicate the trauma outcome measure category by comparing total TEFS at zero weeks versus total TOMS at 12 weeks.)

| | |
|---|---|
| Glasgow Outcome Score (GOS) at 12 weeks | ◯ Death (clinically confirmed death)<br>◯ Persistent vegetative state (severe damage with prolonged state of unresponsiveness and lack of higher mental functions)<br>◯ Severe disability (Severe injury with permanent need for help with daily living)<br>◯ Moderate disability (No need for assistance in everyday life, employment is possible but may require special equipment)<br>◯ Good recovery (Minimal injury with minor neurological and psychological deficits, patient is independent and employable)<br>◯ Lost to follow-up<br>(If patient sustained head injury, please indicate their Glasgow outcome score at the end of 12 weeks follow-up) |
| Glasgow coma numerical score | ________________________ |
| Glasgow Coma Outcome Scale Category | ◯ Favorable outcome (GOS = 4 OR 5)<br>◯ Unfavorable outcome (GOS = 3, 2, OR 1)<br>(Indicate the Glasgow Coma Outcome Scale category) |
| Mode of follow-up | ◯ Through out-patient clinic visit<br>◯ By phone call<br>◯ Home visit<br>◯ In hospital wards and emergency departments<br>◯ Retrieval of hospital records and mortality audits<br>◯ Combination of two or more of the above<br>(How was the patient followed-up?) |
