## Supplemental Material 5 for "Effect of rural trauma team development on outcomes of motorcycle related injuries: A protocol for a multi-center cluster randomized controlled clinical trial (The MOTOR trial)"

**Supplemental material 5: Trainee participant data collection tool**

**PARTS 1 & 2 PRE-AND POST-RAINING QUESTIONNAIRE**

Reg. No…………………………………..Cadre/designation…………………………………….

Hospital where rotation/attachment is held……………………………………………………..

Instructions: (Circle the right answer for EACH of the MCQs 1-20)

1. When an injury occurs in a rural environment, there are many things that may cause a delay in the patient receiving definitive care. These delays result in an increase in preventable morbidity and mortality. Which of the following is NOT a potential delay that should be minimized in the rural environment?
a) Delay in the identification of immediately life-threatening injuries
b) Delay in the treatment prioritization of potentially life-threatening injuries
c) Delay in response time of the urban referral center
d) Delay in transport to higher level of care
2. Which of the following is INCORRECT?
a) The most important determinant of quality care is a timely, organized, rational response to the care of the trauma patient
b) Definitive care may not be possible or desirable in the rural environment
c) If definitive care is NOT possible, an attempt at diagnosing all injuries is not helpful if it results in a delay in transfer to an appropriate level of care
d) Pharmacy, blood bank, and radiology support services are always available in the rural environment
3. A 65-year-old woman is being brought to the emergency department (ED) of a rural facility by Emergency Medical Services (EMS) following a fall down a flight of stairs. Upon arrival, she has a seizure and then becomes unresponsive. To be most effective, the team leader in this situation must do all of the following EXCEPT:
a. Assign tasks/responsibilities
b. Articulate the plan
c. Personally execute all aspects of the plan
d. Adapt the plan on changes in the situation
4. A two-year-old (13 kg) girl is in the ED following a fall through a second-story screen window. She has normal vital signs (age appropriate) but has obvious deformities of both forearms. Prior to reducing and splinting these fractures, the team leader has asked that that patient be given 4 mg of morphine and 2 mg of midazolam intravenously. The nurse is concerned that these amounts may be too much medication. The nurse should:
a) Give the medication as ordered and carefully monitor the patient’s respiratory status
b) Clearly articulate the concern to the team leader in a respectful fashion
c) Report the medication error to the nursing supervisor
d) Secretly give half of the ordered amount and observe the patient’s reaction
5. Absolute indications for intubation include all the following EXCEPT:
a) T3 spinal cord injury
b) Glasgow Coma Score (GCS) of 8 or less
c) Severe maxillofacial injury
d) Neck injury with soft tissue swelling and airway compromise
6. Rapid sequence intubation:
a) Should be done only after the cervical spine is cleared of injury
b) Uses a long-acting paralytic such a pancuronium so that many attempts are possible
c) Is used to help assist with planned placement of a rescue airway device
d) Should be done following initial airway management with a bag-valve mask to allow preoxygenation
7. An 85-year-old female is transferred by ambulance to a 25-bed critical care access hospital with no surgical capability. The patient was found by a neighbor near a mailbox after she reportedly fell on a patch of ice. The exact time the patient was down on the ground is unknown. She is unresponsive to verbal and painful stimuli. Her heart rate is 50, blood pressure is 170/80, and she has respirations of 16. The nearest trauma center with neurosurgical capability is one hour away by fixed-wing aircraft. Prior to transfer, how should the airway be managed?
a. Does not need any airway intervention
b. Endotracheal intubation
c. Cricothyroidotomy
d. Supplemental oxygen using nasal cannula
8. All of the following injuries must be identified and immediately addressed during the primary survey EXCEPT:
a) Simple pneumothorax
b) Massive hemothorax
c) Tension pneumothorax
d) Open pneumothorax
9. A 25-year-old male was involved in a high speed, single vehicle crash. He is brought to a 25-bed, critical care access hospital with no surgical capability. His blood pressure is 70/30, heart rate is 135, respiratory rate is 45, and he is unresponsive to verbal stimuli. An emergency medical technician (EMT) is manually ventilating the patient with a bag valve mask. The patient’s neck veins are flat with no associated tracheal deviation. Initial auscultation of the chest reveals absent breath sounds on the right along with dullness to percussion. There is no obvious penetrating trauma. The diagnosis related to the thorax is:
a) Open pneumothorax
b) Flail chest
c) Tension pneumothorax
d) Massive hemothorax
10. A tension pneumothorax may be rapidly fatal. Clinical signs and symptoms of this include all of the following EXCEPT:
a) Muffled heart tones
b) Respiratory distress
c) Distended neck veins
d) Unilateral absence of breath sounds
11. The most common cause of shock in the injured patient is:
a) Cardiogenic
b) Hemorrhagic
c) Neurogenic
d) Septic
12. A 40-year-old man was involved in a motorcycle crash in which he was not wearing a helmet. His GCS is 3, heart rate is 120, and blood pressure is 90/60. There is external blood loss from his scalp. The volume of blood loss to cause shock in this adult patient can be found in all of the following locations EXCEPT:
a) External hemorrhage
b) Intrathoracic hemorrhage
c) Pelvic hemorrhage
d) Intracranial hemorrhage
13. Which of the following is TRUE regarding compensated shock?
a) Cardiac output is normal
b) Organ perfusion and tissue oxygenation is decreased
c) Hypotension is always present
d) Tachycardia is a sensitive sign in all elderly patients
14. Providers at a rural hospital without surgical resources must:
a) Determine the cause of shock before initiating the transfer process
b) Repair all lacerations before transfer
c) Obtain radiographs and treat all fractures before transfer
d) Initiate the transfer process when you suspect the patient has injuries that exceed your local resources
15. Secondary brain injury:
a) Refers to damage that occur shortly after injury
b) Does not influence overall outcome
c) Can be minimized by maintaining oxygenation and blood pressure
d) Is the same as spinal shock
16. A 32-year-old woman has a GCS of 8 after a fall down a flight of stairs. Her blood pressure is 85/60 and heart rate is 120. Her treatment must include:
a) Hypotonic fluid resuscitation
b) Mannitol
c) Hyperventilation
d) Endotracheal intubation
17. It is a warm summer afternoon. A 58-year-old male pedestrian was struck by a motor vehicle an hour ago and is now being assessed in your rural hospital emergency department. He is tachycardic with a blood pressure of 90/60, is restless and somewhat uncooperative, and is being resuscitated with crystalloids. Which of the following statements is correct?
a) Hypothermia is not a significant concern in this patient
b) Oxygen saturation measurements will be highly reliable in this individual
c) Room temperature should be optimized for trauma team members’ efficiency and comfort
d) Oral temperature measurements in this patient may not be reliable
18. You are assessing a 24-year-old woman in your community hospital emergency department. She was thrown from her snowmobile. Your team suspects a closed head injury, pelvic trauma, and a fractured femur. All of the following statements regarding her resuscitation are true EXCEPT:
a) The patient’s wet clothing should be removed
b) The receiving area should be warmed to 80 degrees Fahrenheit (27 degrees Celsius) prior to her arrival
c) Packed red cells should be placed in a microwave oven briefly before administration
d) Passive rewarming (dry blankets, Mylar sheets, plastic wrap) should be used
19. A Broselow Tape is:
a) Used to secure an endotracheal tube
b) A special adhesive used to secure cricothyroidotomy tube
c) Used to determine pediatric drug dosing
d) Used to assure inline stabilization of the neck for intubation
20. Children are less prone to rib fractures and mediastinal trauma and more prone to pulmonary parenchymal injuries because:
a) Children are rarely involved in traumatic incidents
b) Their lungs are less compliant and do not resist injury as well
c) Children have a compliant chest wall and mobile mediastinum
d) Children’s ribs are stronger and therefore resist fracture

The pre and post course questionnaire is with permission, based on educational materials of rural trauma team development course 4^th^ edition of the American College of Surgeons

**PART 3: BARRIERS TO INJURY CARE**

**Please rank the following delays as barriers to timely provision of definitive injury care for trauma patients in general. For each question, please use each numeric number once.**

1. In your current hospital of attachment, please rank the potential significant **system delays** to providing care to the injured (trauma) patients, in order of importance from the most important (score of 6) to the least important (score of 1) by assigning numbers 1, 2, 3, 4, 5, 6
a. Delays due to Discovery of the injured in the field
b. Delays in summoning/calling for help
c. Delays in mobilizing emergency medical service (EMS)/ambulance providers
d. Delays due to pre-hospital response time (ambulance arrival to scene)
e. Prolonged scene time (Delay at the scene)
f. Long transportation time to the hospital
g. Other (specify)……………………………………………………………………………
2. After the trauma patient has arrived to your current hospital of attachment, please rank the potential significant **team delays** to providing care to such patient in order of importance (by assigning a score of 4 to the most significant and 1 to the least significant (use 1,2,3,4)
a. Delays in identification of immediately life-threatening injuries
b. Delays in prioritization of life-threatening injuries
c. Delays in recognition of injury severity
d. Delays in transportation to a higher level of care
e. Other (specify)…………………………………………………………………………
3. In your current hospital, after the decision has been made to carry out emergency surgery for a trauma patient (for example amputation for a crushed limb), please rank the potential significant **barriers** to providing surgery to such patient in order of importance (by assigning a score of 4 to the most significant and 1 to the least significant. (use numbers 1,2,3,4)
a. Lack of skilled staff (surgical or anesthetic) to carry out the surgery
b. Lack of theatre space/functional theatre to carry out the surgery
c. Lack of supplies such as oxygen, emergency drugs, blood products, sutures
d. Lack of intensive and critical care (ICU) services for post-operative management
e. Other (specify)……………………………………………………………… …

**PART 4: PARTICIPANT COURSE EVALUATION FORM**

Instructions: This form provides an opportunity for you to rank each of the individual topics by ticking the appropriate box based on your overall individual experience and what you think of this kind of training.

| **S/N** | **Topics** | **Strongly Agree(5)** | **Agree (4)** | **Neutral(3)** | **Disagree (2)** | **Strongly Disagree (1)** |
| --- | --- | --- | --- | --- | --- | --- |
| 1 | Overall, this educational activity was excellent |  |  |  |  |  |
| 2 | Program topics and content met the stated objectives |  |  |  |  |  |
| 3 | Content was relevant to my educational needs |  |  |  |  |  |
| 4 | Educational format was conducive to learning |  |  |  |  |  |
| 5 | Acquired knowledge will be applied in my practice environment |  |  |  |  |  |
| 6 | I will seek additional information on this subject |  |  |  |  |  |
| 7 | Program was fair, objective, and unbiased towards any product or program |  |  |  |  |  |
| 8 | Power point slides are well written, visually appealing with good references |  |  |  |  |  |
| 9 | The audiovisuals enhance the presentation |  |  |  |  |  |
| 10 | Course format (lecture/ skill station scenarios) stimulates critical thinking |  |  |  |  |  |
| 11 | Content is organized in a concise and logical sequence |  |  |  |  |  |
| 12 | Instructor has knowledge about content |  |  |  |  |  |
| 13 | Instructor presentation style keeps learner’s attention |  |  |  |  |  |
| 14 | Instructor uses examples to illustrate major points |  |  |  |  |  |
| 15 | Instructor presents content accurately and confidently |  |  |  |  |  |
| 16 | Instructor answers questions in a supportive manner |  |  |  |  |  |
| **For each of the performance improvement/patient safety, communication & scenarios, rank the relevance of each module by ticking the corresponding box in the shaded area** | | | | | | |
| Rank (score) | | Performance improvement & patient safety | | Communication module | | Case scenarios |
| 1 | Not relevant |  | |  | |  |
| 2 | Relevant |  | |  | |  |
| 3 | Very relevant |  | |  | |  |

Adapted with permission from: Ali J, Kumar S, Gautam S, Sorvari A, Misra MC. Improving Trauma Care in India: the Potential Role of the Rural Trauma Team Development Course (RTTDC). Indian J Surg. 2015;77:227–31.
