## Supplementary material for "Effect of rural trauma team development on outcomes of motorcycle related injuries: A protocol for a multi-center cluster randomized controlled clinical trial (The MOTOR trial)": SPIRIT Checklist

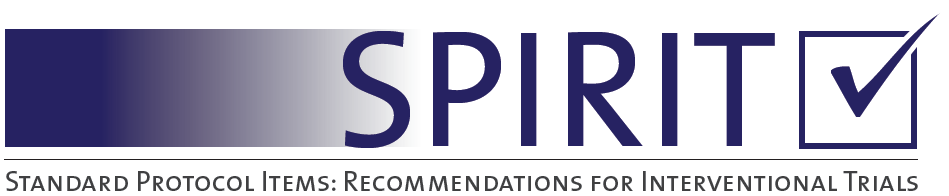

SPIRIT 2013 Checklist: Recommended items to address in a clinical trial protocol and related documents*

| Section/item | | Item No | | | Description | Pg | |
| --- | --- | --- | --- | --- | --- | --- | --- |
| **Administrative information** | | | | | |  | |
| Title | | 1 | | | Effect of rural trauma team development on outcomes of motorcycle related injuries: A protocol for a multi-centre cluster randomized controlled clinical trial (The MOTOR trial) | 1 | |
| Trial registration | | 2a | | | Pan African Clinical Trial Registry (PACTR202308851460352) | 2 | |
|  |  | 2b | | | Since the country where the trial was approved did not have a publicly available registry, the protocol was retrospectively registered with WHO approved Pan African Clinical Trials Registry. | 2 | |
| Protocol version | | 3 | | | Protocol version V November 2023 | 3 | |
| Funding | | 4 | | | This study did not receive any external funding. HL was enhanced with a research loan from the Uganda Medical Association to support this study. JPP is supported by the Academy of Finland (grant no 17379) and the Maire Taponen Foundation. The sponsors did not have any role in the design, writing or decision to submit the protocol for publication. | 3 | |
| Roles and responsibilities | | 5a | | | HL is the principal investigator. MAM is study biostatistician, RS and PK are local study overseers, TB, JPP and MLW are overseas study supervisors. | 3 | |
|  |  | 5b | | | Herman Lule; | 1 | |
|  | | 5c | | | This trial has not received external funding | 3 | |
|  | | 5d | | | The principal investigator and a central study administrator will form a steering committee that will run the day-to-day activities of the trial. The trial will be overseen by two onsite and two off-site supervisors. In addition, the principal investigator will provide organizational support to rural trauma teams at the intervention study centres through weekly audit meetings | 22 | |
| Introduction | |  | | |  |  | |
| Background and rationale | | 6a | | | Injury is a global health concern whose mortality disproportionately impact low-income countries. Compelling evidence from high-income countries show that rural trauma team development courses (RTTDC) increase clinicians’ knowledge. There is a dearth of evidence from controlled clinical trials to demonstrate the effect of RTTDC on process and patient outcomes. We document a protocol for a multi-centre cluster randomized controlled clinical trial which aims to examine the impact of RTTDC on process and patient outcomes of motorcycle-related injuries. | 3-5 | |
|  | | 6b | | | This will be a two-armed parallel multiple period cluster randomized controlled clinical trial in Uganda, where rural trauma team development training is not routinely conducted. We shall recruit regional referral hospitals and include road traffic injured patients, interns, medical trainees, and road traffic law enforcement professionals who serve as trauma care frontliners. Three hospitals will be randomized to RTTDC (intervention group) and the remaining three to standard care (control group). | 4,5 | |
| Objectives | | 7 | | | I. To determine the effect of rural trauma team development course training on crash scene-admission interval and outcome of musculoskeletal injuries. II. To determine the effect of rural trauma team development course training on the referral decision to hospital exit interval and outcome of neurological injuries. | 5,6 | |
| Trial design | | 8 | | | This will be a two-armed parallel multiple period cluster randomized controlled clinical trial | 6 | |
| Methods: Participants, interventions, and outcomes | | | | | |  | |
| Study setting | | 9 | | | This study will be conducted in six specialized teaching and referral hospitals in Uganda where RTTDC is not taught, including: Kiryandongo, Jinja, Hoima, Fort portal, Mubende and Kampala International University Hospital. These facilities have similar characteristics by serving as teaching, residency and internship sites for undergraduate, graduate medical doctors and nurses. The hospitals offer 24/7 emergency surgical services for trauma patients through multidisciplinary teams. Each surgical department in these facilities is typically composed of about 1-4 faculty, 2-4 specialty residents, 4-6 interns and 10-30 undergraduate medical trainees. | 6,7 | |
| Eligibility criteria | | 10 | | | **Inclusion criteria**  i. Study sites: Level 3 trauma centres which are teaching hospitals, staffed with medical trainees, interns, surgery residents and consultants; offering 24/7 emergency surgical care with access to blood banks, ultrasound, X-ray, and CT scan as locally available or as outsourced services.  ii. Trainee participants: Third year or fifth year medical or allied health students, interns or specialty residents in surgery and traumatology clinical rotations who will stay at the hospital of attachment for at least two-three months. In addition, we shall include road traffic law enforcement professionals concerned with evacuation of trauma patients from accident scenes.  iii. Patient participants: Patients who sustain motorcycle-related injuries and present to study sites within 24 hours following crash as defined in previous studies. These will include passengers on motorcycles, motorcycle riders, pedestrians hit by motorcycles, patient suffering from of motorcycle-motorcycle collisions or motorcycle-static object collisions, passengers on a motorcycle or cyclists suffering from motorcycle-car collisions.  **Exclusion criteria**  i. Trauma centres which do not offer placements and teaching facilities for students, interns, and residents.  ii. Trainee participants: Medical students who have not commenced their surgery clinical rotation and those not directly involved with care of trauma patients at the time of the training since the trainee medical participants are expected to have already been introduced to surgery, emergency trauma resuscitation concepts and trauma clinical scenarios through clerkships onto which we shall build the rural trauma team concept.  iii. Patient participants: Pregnant women, neonates, and infants 0-23 months will be excluded due to the known teratogenic effects of radiations to this population since trauma evaluation in this study will involve obtaining radiographs for orthopaedic injuries and Computerized Tomographic (CT) scans for neurological injuries. Patients with documented stroke will also be excluded since the study outcomes involve assessment for functional, physical, and neurological disability directly attributable to trauma. Mentally incapacitated patients who have no legally authorized representatives to sign an informed consent will be excluded as well as patients who die before hospital arrival or before imaging results are obtained. Patients who are passengers in a car or are drivers in a car at the time of crash will be excluded given the “protective casing of the car body” which would make these patients less vulnerable to direct impacts compared to pedestrians, cyclists, or passengers on motorcycles. In addition, elderly over 80 years will be excluded due to their increased risk of fragility fractures that could misrepresent the severity of trauma. | 7, 8 | |
| Interventions | | 11a | | | The 2 day RTTDC interventional training will be conducted in designated spacious multimedia surgical simulation conference rooms at the study sites randomized to intervention arm, and will be delivered in its described standard format (see reference).  Ref: The American College of Surgeons. Rural Trauma Team Development Course [Internet]. Trauma Education. [cited 2023 Aug 30]. Available from: <https://www.facs.org/quality-programs/trauma/education/rural-trauma-team-development-course/> | 9 | |
|  |  | 11b | | | The 4th edition of RTTDC will be delivered in its standardized form without any modifications. | 10 | |
|  |  | 11c | | | RTTDC is the intervention thus participants who will not be able to attend all the modules and complete the post training MCQs will be excluded. Adherence to RTTDC teachings amongst care providers during their clinical practice such as time taken from referral decision to execution will be assessed as process measure outcomes. | 10 | |
|  |  | 11d | | | The baseline quality of care given to trauma patients at each of the participating six hospitals is based on the regulations of the Uganda Medical and Dental Practitioners Council (UMDPC) and the National Council for Higher Education (NCHE) which regulate both undergraduate and graduate surgical curriculum. In terms of standards of care, both intervention and control facilities have medical trainees, interns, specialty residents and specialists who provide care for injured patients. The care typically involves receiving trauma patients who may be brought by staffed ambulances, police or public means to the accident and emergency units, and are received by trainees, interns, or surgical residents. These cadres open case files, initiate immediate care through horizontal consultations, request X-rays, CT scans and laboratory workup, plan definitive care or referrals in liaison with a third-year senior surgical resident, and in consultation with a multidisciplinary team of specialists on call. | 10, 11 | |
| Outcomes | | 12 | | | The primary outcomes will be prehospital interval from accident scene to arrival at emergency department, and referral-exit interval from the time the referral decision is made to hospital exist in hours as a measure of process improvement.  The secondary outcomes will be all cause mortality, and morbidity of neurological, and orthopaedic injuries based on the Glasgow outcome scale and trauma outcome measure scores respectively at 90-days post injury. All outcomes will be measured as final values.  The tertiary outcomes will be the effect of the training on providers knowledge based on pre-and post-training trauma related MCQ scores. | 12 | |
| Participant timeline | | 13 | | | see SPIRIT Figure 2 attached | 13 | |
| Sample size | | 14 | | | We estimated a total sample size of 501 per treatment arm using open-source R shiny application for cluster randomized controlled trials with parallel design and discrete time decay correlation structures for multiple periods available at <https://clusterrcts.shinyapps.io/rshinyapp/> | 13, 14 | |
| Recruitment | | 15 | | | Course participants will be recruited at the surgical simulation conference rooms by advertisement through hospital administrators, class representatives and regional traffic law enforcement leaders. Patient participants will be recruited at the respective accident and emergency departments. | 15 | |
| **Methods: Assignment of interventions (for controlled trials)** | | | | | |  | |
| Allocation: | | 16 | | | Trauma centres will be assigned to either intervention (RTTDC) group or control (standard care) group. | 15 | |
| Sequence generation | | 16a | | | We shall perform cluster randomization for a permuted block size of six hospitals (clusters) using an open-source simulation software available at (https://www.sealedenvelope.com/simple-randomiser/v1/lists), with seed numbers that will be kept confidential to the offsite study administrator. A list length of six random codes will be generated to assign hospitals as intervention or control groups. | 15 | |
| Allocation concealment mechanism | | 16b | | | The allocation sequence and assignment codes will be generated by and kept secret by an offsite study administrator. | 15 | |
| Implementation | | 16c | | | Trainee participants will be enrolled by the respective university hospital administrators whereas patient participants will be enrolled by project medical officers at the respective study sites who will also serve as outcome assessors. | 15, 16 | |
| Blinding (masking) | | 17a | | | Both outcome assessors and patient participants will be blinded by the treatment allocation. | 16 | |
|  | | 17b | | | Unblinding will only be permissible in the likely adverse event to the study participants that can be directly attributable to the study intervention. Such events will be discussed with the attending physicians. | 16 | |
| **Methods: Data collection, management, and analysis** | | | | | |  | |
| Data collection methods | | 18a | | | T Data collection will start simultaneously at all the study sites. Medical officers (blinded outcome assessors) with a minimum qualification of Bachelor of Medicine and Bachelor of Surgery (MBChB) and at least two years of clinical experience will prospectively collect data using pre-designated questionnaires through clinical observations, clinical interviews, home visits, and extraction from hospital and police records. The data collection tools are available as supplementary materials 4 and 5. | 16 | |
|  | | 18b | | | Phone contacts for the patients, their legally authorized representatives or local village chairpersons will be obtained. In return, study participants will be availed with the phone contacts of study administrator, principal investigator, and dedicated study nurse coordinators who will make reminder calls to participants due for appointment to minimize loss to follow-up. | 17 | |
| Data management | | 19 | | | Data will be collected on hard copies, transferred to Research Electronic Data Capture (REDCap) secure virtual network hosted by the University of Turku, and later exported to Stata 15.0 for analysis. | 18 | |
|  | Statistical methods | | 20a | Primary outcomes:  The mean (i) pre-hospital interval and (ii) referral-exit interval in hours (standard deviation) between intervention and control groups will be compared using a two-sample t-test which is robust for normality deviations.  Secondary outcomes:   (i) The difference in proportions of all-cause mortality at 90 days between the intervention and control arms will be compared using Chi-square test if the expected counts are >5, otherwise by use of Fisher’s exact test at 95%CI. (ii) The mean difference in GOS will be compared between groups using the two sample t-test . Further, the GOS will be dichotomized into favorable (GOS=4 or 5) and unfavorable (GOS=3 or 2 or 1) outcomes. The difference in proportions between the intervention and control group will be compared using adjusted Chi-square test if the expected counts are >5, otherwise by use of Fisher’s exact test at 95%CI. (iii) The mean difference in TEFS and TOMS will be compared between groups using the two sample t-test. Further, the TOMS will be dichotomized into favorable (TOMS≥TEFS) and unfavorable (TOMS<TEFS) outcomes and the difference in these proportions between the intervention and control groups will be compared using adjusted Chi-square test if the expected counts are >5, otherwise by use of Fisher’s exact test at 95%CI.   Tertiary outcomes (Ancillary studies):   (i) The difference in pre- and post-training mean scores or median and interquartile range will be computed at 95%CI. (ii) For perceived barriers to injury care, directed content analysis of themes of transcribed data will be collated manually and presented as percentages | | | 19 |
|  | | 20b | | | Subgroup analyses will be performed for: (i) varying road user categories (pedestrian, passenger, motorcyclists), (ii) injury mechanisms (motorcycle-motorcycle crash, motorcycle-pedestrian crash, motorcycle-static object, motorcycle-car crash), (iii) varying injury severities (mild, moderate, or severe based on the Kampala Trauma Score and Glasgow Comma Scale), (iv) multiplicity of serious injuries (one or multiple) and (v) study time periods. | 21, 22 | |
|  | | 20c | | | We shall impute values for participants with missing end points such as lost to follow-up, withdrew consent, and crossovers. The baseline sociodemographic and clinical characteristics of such participants will be compared between the two treatment groups (intervention vs. control) and to those retained within the treatment groups. For individual participants, crossover from intervention to control group or vice versa due to any reason will result in ultimate discontinuation from the trial. | 21 | |
| **Methods: Monitoring** | | | | | |  | |
| Data monitoring | | 21a | | | The Research and Ethics Committee of Mbarara University of Science and Technology is the data monitoring committee for this trial under Ref (MUREC 1/7; 05/5-19). The committee reports directly to Uganda National Council for Science and Technology. | 22 | |
|  | | 21b | | | The Uganda National Council for Science and Technology accessed a preliminary report of interim analyses which were performed during 28 August 2022 when the half of the sample size was attained and at its discretion recommended continuation of the study.  Recommendation to terminate an individual participant from the trial will be made by the principal investigator in the event of breach of trial protocol such as cross over or loss of follow-up. | 21 | |
| Harms | | 22 | | | Immediate medical concerns from participants will be reported to their attending clinicians. Any reported adverse events and other unintended effects of trial interventions or trial conduct will be reported to the trial monitoring committee. | 22 | |
| Auditing | | 23 | | | The trial monitoring committees will independently conduct annual and impromptu audits and may choose to extend or terminate the trial at any time. Such situations that could warranty termination include adverse events directly attributable to the study or the unlikely event of the trial achieving a meaningful difference in study outcomes between groups. | 22 | |
| Ethics and dissemination | | | | | |  | |
| Research ethics approval | | 24 | | | This study was approved and registered by the research and ethical committee of Uganda National Council for Science and Technology (Ref. No. SS 5082) prior recruitment. Written informed consent will be obtained from all study participants. | 26 | |
| Protocol amendments | | 25 | | | Any protocol amendments will be submitted to the data monitoring committee. Any approved amendments will be communicated to Uganda National Council for Science and Technology, trial participants, and updated in the registries within five working days following approval. | 22 | |
| Consent or assent | | 26a | | | Written informed consent will be obtained for both trainee and patient participants prior to participation. The participants or their legally authorized representatives (for minors and unconscious) will endorse a predesignated consent form document with their signatures in the presence of principal investigator or research assistants (surgery specialty residents). | 8 | |
| Specimen handling | | 26b | | | There will be no biological specimens retained for this study. Informed consent form documents will have provisions for consenting to follow-up, use of data for approved ancillary studies and for permission to archive the anonymized data on a public data repository. | 8 | |
| Confidentiality | | 27 | | | To protect participants’ confidentiality before, during, and after the trial, all hard copy data collection items will be kept under lock and key and will bear unique non identifying codes. Soft copies will be kept in password protected computers with a second layer protection login password required for Redcap, only accessible by the study team. The final data sets will be anonymized prior to archive and publication. | 19 | |
| Declaration of interests | | 28 | | | All authors declare no competing interest. | 26 | |
| Access to data | | 29 | | | Data will be kept in password protected files only known to the investigators until final stages of dissemination. The anonymized datasets arising from this study will be archived and made publicly available through a permanent weblink to a repository that will be provided by the publishing peer reviewed journal. Since the primary country of recruitment which approved and registered the study prior recruitment lacked a publicly available electronic research register, the full protocol was retrospectively registered with WHO approved Pan African Clinical Trial Registry (PACTR202308851460352). | 26 | |
| Ancillary and post-trial care | | 30 | | | Any participants requiring further care beyond the 90-day follow-up will be linked to their attending clinicians during an additional three-months period after completion of the trial. | 11 | |
| Dissemination policy | | 31a | | | Participants for the RTTDC training will get feedback after the post-test questionnaire evaluation. Patient participants will be advised on the next plans of management during follow-up calls or through out-patient consultations. Before study findings will be presented in scientific conferences and published in peer reviewed journals, a copy of findings in the final bound report will be submitted to the participating hospital main libraries, departments of surgery and Internal Review Boards of Mbarara University of Science and Technology, Kampala International University and Uganda National Council for Science and Technology. The implications of findings will be shared with to authorities including interns and medical students’ associations, district health officers, hospital directors, regional traffic police commanders and chairpersons of the motorcycle riders' associations in the respective regions, and with the rural trauma networks of first responders. | 23 | |
|  | | 31b | | | HL Principal investigator, Conceptualization, Data curation, Investigation, Methodology, Project administration, Resources, Writing-original draft; HL, MM Formal analysis, Software, Visualization; RS, PK, TB, JPP, MLW Validation, Writing-review, and editing; TB, JPP, MLW Supervision, Funding acquisition. All authors read and approved the final manuscript. | 25 | |
|  | | 31c | | | The full protocol will be published open access and anonymized participant-level datasets, and statistical codes will be shared publicly through a permanent weblink that will be provided by the publishing journal. | 21 | |
| Appendices | |  | | |  |  | |
| Informed consent materials | | 32 | | | Model consent forms for trainee and patient participants are available as online supplementary material 1 and 2 respectively | 26 | |
| Biological specimens | | 33 | | | There are no plans to collect any biological laboratory specimens either for purposes of storage, genetic or molecular analysis in this trial. | 19 | |

*It is strongly recommended that this checklist be read in conjunction with the SPIRIT 2013 Explanation & Elaboration for important clarification on the items. The SPIRIT checklist is copyrighted by the SPIRIT Group under the Creative Commons “[Attribution-Non-commercial-NoDerivs 3.0 Unported](http://www.creativecommons.org/licenses/by-nc-nd/3.0/)” license.
