## Supplementary material for "Effect of rural trauma team development on outcomes of motorcycle related injuries: A protocol for a multi-center cluster randomized controlled clinical trial (The MOTOR trial)": Ethical Approval

### Uganda National Council for Science and Technology

*(Established by Act of Parliament of the Republic of Uganda)*

UNCST

**Our Ref: SS 5082** **2nd September 2019**

Dr. Herman Lule
Principal Investigator
Kiryandongo General Hospital
**Kigumba**

Dear Dr. Lule,

**Re: Research Approval: The Use of Allied Health and Law Enforcement Trauma Team Registries: Lessons from a Novel Motorcycle Trauma Registry and Surveillance System in Uganda**

I am pleased to inform you that on **27/08/2019**, the Uganda National Council for Science and Technology (UNCST) approved the above referenced research project. The Approval of the research project is for the period of **27/08/2019** to **27/08/2023.**

Your research registration number with the UNCST is **SS 5082.** Please, cite this number in all your future correspondences with UNCST in respect of the above research project.

As Principal Investigator of the research project, you are responsible for fulfilling the following requirements of approval:

1. All co-investigators must be kept informed of the status of the research.
2. Changes, amendments, and addenda to the research protocol or the consent form (where applicable) must be submitted to the designated Research Ethics Committee (REC) or Lead Agency for re-review and approval **prior** to the activation of the changes. UNCST must be notified of the approved changes within five working days.
3. For clinical trials, all serious adverse events must be reported promptly to the designated local IRC for review with copies to the National Drug Authority.
4. Unanticipated problems involving risks to research subjects/participants or other must be reported promptly to the UNCST. New information that becomes available which could change the risk/benefit ratio must be submitted promptly for UNCST notification after review by the REC.
5. Only approved study procedures are to be implemented. The UNCST may conduct impromptu audits of all study records.

6. An annual progress report and approval letter of continuation from the REC must be submitted electronically to UNCST. Failure to do so may result in termination of the research project.

**Below is a list of documents approved with this application:**

| | Document Title | Language | Version | Version Date |
|---|---|---|---|---|
| 1. | Research proposal | English | N/A | N/A |
| 2. | Informed consent forms | English and Runyankole | N/A | N/A |
| 3. | Investigator administered questionnaire | Runyankole | N/A | N/A |
| 4. | PRISMA flow chart for article selection | English | N/A | N/A |
| 5. | Systematic review check list of included articles | English | N/A | N/A |
| 6. | Pre and post questionnaire | English | N/A | N/A |
| 7. | Barriers to injury care | English | N/A | N/A |
| 8. | Participant course evaluation form | English | N/A | N/A |
| 9. | Motorcycle trauma and outcome registry data sheet | English | N/A | N/A |
| 10. | Trauma Expectation Factor Score (TEFS) | English | N/A | N/A |
| 11. | Trauma Outcome Measure Score (TOMS) | English | N/A | N/A |
| 12. | Glasgow outcome scale | English and Runyankole | N/A | N/A |

Yours sincerely,

Isaac Makhuwa
For: Executive Secretary
**UGANDA NATIONAL COUNCIL FOR SCIENCE AND TECHNOLOGY**

Copied to: Chair, Mbarara University of Science and Technology, Research Ethics Committee
